# Generalizability of EEG-Based Dementia Classifiers: A Multicenter study of Alzheimer’s, MCI, And FTD

**DOI:** 10.64898/2026.07.28.26359133

**Authors:** Laouen Belloli, Nicolás Bruno, Hernan Hernandez, Jhosmary Cuadros, Damián Dellavale, Pavel Prado, Renato Anghinah, Bahar Güntekin, Lütfü Hanoǧlu, Mario A Parra, Agustín Ibañez, Jacobo Sitt

## Abstract

EEG-based machine learning shows promise for neurodegenerative disease classification, but robustness to sample imbalance, center heterogeneity, and validation leakage remains a key concern for clinical translation. We developed a new framework to assess diagnostic performance, calibration, and cross-center generalizability of EEG multifeatured classifiers across CN (cognitively normal), MCI (mild cognitive impairment), AD (Alzheimer’s disease), and FTD (frontotemporal dementia), while addressing imbalance, statistical uncertainty, and validation rigor across six centers. Supervised classifiers were evaluated at aggregated- and subject-level repeated cross-validation and leave-one-center-out (LOCO) schemes, and calibration was implemented via Platt scaling within strictly nested folds. CN vs AD classification showed robust performance and cross-center generalizability, with consistent AUC and calibration across cross-validation and leave-one-center-out analyses. In contrast, CN versus MCI showed moderate, heterogeneous performance and limited cross-center generalizability, with chance-level results in some cohorts, while MCI versus AD showed moderate discrimination in a single available center. FTD contrasts showed modest or limited performance due to sparse samples. Predicted probabilities were stable across validation regimes for AD, but less consistent for MCI and FTD, and correlated robustly with cognitive impairment severity only for AD. Feature importance analyses identified disease-specific signatures, including alpha-band degradation and slow-wave increases in AD, with weaker and more heterogeneous patterns in prodromal and differential dementia contrasts (FTD vs AD). EEG classifiers provided robust discrimination for CN vs AD but showed limited and heterogeneous performance for MCI and FTD across centers. These results emphasize the need for balanced sampling, strict validation of clinical and EEG protocols, and uncertainty quantification to support reliable clinical deployment.

## 1 Introduction

Neurodegenerative dementias represent one of the most pressing healthcare challenges of the 21st century, with a substantial increase in projected global prevalence (GBD 2019 Dementia Forecasting Collaborators, 2022; Custodio et al., 2017; Livingston et al., 2020). This demographic transition disproportionately affects low- and middle-income countries, where exposure to risk factors including socioeconomic disparities, environmental pollutants, and infectious diseases accelerates brain aging (Legaz et al., 2025; Hernandez et al., 2025; Prado, Medel, et al., 2023). The implementation of early and affordable diagnostic strategies is vital in these settings, as timely detection provides significant health-economic benefits and improves clinical outcomes (Kwon et al., 2025). Mainstream diagnostic frameworks have traditionally relied on pathological biomarkers such as amyloid beta and tau proteins, quantified through positron emission tomography (PET) or cerebrospinal fluid (CSF) analysis (Jack et al., 2018; McKhann et al., 2011; Dubois et al., 2014). While these methods offer high sensitivity, their clinical utility in global settings is hindered by prohibitive costs and extremely limited access, particularly in developing economies (Parra et al., 2021; Parra et al., 2023; Custodio et al., 2024; Duran-Aniotz et al., 2021). Although blood-based (plasma) biomarkers have emerged as a promising alternative (Ntymenou et al., 2021), they are not yet widely accessible and lack systematic validation in populations (Parra et al., 2018; Coronel-Oliveros et al., 2024). Moreover, the accuracy of these markers in diverse populations can be influenced by whole-body health (Ibáñez et al., 2025), and usually requires the combination of neuroimaging and cognitive assessments (Caviedes et al., 2026).

Electroencephalography (EEG) emerges as a cost-effective complement, also providing a rich characterization of temporal brain dynamics (Rossini et al., 2020). EEG is uniquely scalable for global initiatives, being non-invasive, portable, low-cost, and widely available across clinical settings (Whelan et al., 2022). This technique has identified multiple neurophysiological signatures of neurodegeneration, such as spectral slowing, reduced signal complexity, and network disconnection observed in Alzheimer’s disease (Moguilner et al., 2024; Prado, Mejía, et al., 2023; Babiloni et al., 2021; Dauwels et al., 2011). Furthermore, distinct oscillatory, topographic and mechanistic patterns differentiate dementia subtypes, such as Alzheimer’s, frontotemporal dementia and mild cognitive impairment (Coronel-Oliveros et al., 2025; Jelic et al., 2000; Nishida et al., 2011). These markers can feed automated classification algorithms, demonstrating good diagnostic accuracy by exploiting multivariate patterns (Akbar et al., 2025; Yuan & Zhao, 2025; Moguilner et al., 2022). Thus, computational multifeature EEG architectures can help characterize dementia in global settings.

Despite this progress, several factors could hinder the clinical translation of EEG-based biomarkers. Prior studies are frequently constrained by methodological weaknesses, including small sample sizes, potential data leakage, and insufficient statistical validation, which may inflate performance estimates and undermine reproducibility. Moreover, technical, demographic, and site-specific heterogeneity (Prado et al., 2022) spanning EEG acquisition systems, preprocessing pipelines, and population diversity introduces variance that may obscure disease-specific signals and compromise robustness in diverse cohorts (Moguilner et al., 2024). Existing work usually lacks rigorous multi-center validation using leave-one-center-out or similarly stringent frameworks (Prado, Mejía, et al., 2023; Peh et al., 2021), leaving generalizability largely unproven. Beyond technical performance, most approaches remain poorly aligned with clinical decision-making, relying on binary classifications rather than calibrated probabilistic outputs anchored in neuropsychological standards. These approaches often lack interpretability regarding the neurophysiological features driving predictions, limiting clinical relevance and regulatory adoption (Malik et al., 2025). Finally, much of the literature focuses narrowly on distinguishing Alzheimer’s disease from cognitively normal controls, with insufficient attention to clinically essential challenges (Odusami et al., 2024) such as differential diagnosis necessary for meaningful clinical deployment (Rossini et al., 2020).

We aimed to bridge these gaps through a systematic multi-center design spanning five independent cohorts across diverse cultural and geographical settings. We implemented a “plug-and-play” automated framework that prioritized automaticity to eliminate subjectivity and enhance generalizability across clinical settings. We included complementary domains (from spectral markers and signal complexity to inter-regional connectivity), to capture the multifaceted nature of neural dysfunction (Sitt et al., 2014; King et al., 2013). Although this framework had been previously applied in dementia research, those studies relied on reduced single-center designs with homogeneous cohorts (Gaubert et al., 2019, 2021), leaving its robustness across heterogeneous, multi-site settings untested. This framework has also been employed successfully in multicentric disorders of consciousness research (Sitt et al., 2014; Engemann et al., 2018; Pérez et al., 2024; Manasova et al., 2024) and sleep research (Strauss et al., 2022; Türker et al., 2023). A key advantage of this framework is its potential to maintain stable predictive accuracy even in low-density channel configurations, which supported its feasibility for resource-limited or purely clinical contexts. To rigorously evaluate model generalization, we implemented a three-tier validation strategy: single-center repeated stratified k-fold cross-validation, pooled multi-center stratified k-fold cross-validation, and leave-one-center-out cross-validation as the gold standard for assessing cross-site robustness. Furthermore, our clinical evaluation extended beyond AD versus CN comparisons to include CN versus MCI for early detection, MCI versus AD for disease stage, and FTD versus AD for differential diagnosis. Finally, we incorporated calibrated probability estimation to generate scores that could be directly interpreted in clinical decision-making (Niculescu-Mizil & Caruana, 2005), alongside SHAP-based feature importance to identify center-independent EEG biomarkers with the highest relevance (Figure 1). We hypothesized that multifeature EEG classifiers would demonstrate significant cross-center generalizability despite heterogeneous acquisition conditions, with performance decreasing as validation stringency increases. A graded discriminative pattern should present highest accuracy for CN vs AD, intermediate performance for MCI-related contrasts, and lower accuracy for differential diagnosis between dementia syndromes due to greater neurophysiological overlap. EEG-derived probabilistic predictions would show clinically meaningful associations with cognitive impairment and rely on physiologically interpretable biomarkers consistent with known neurodegenerative mechanisms.

**Figure 1:**
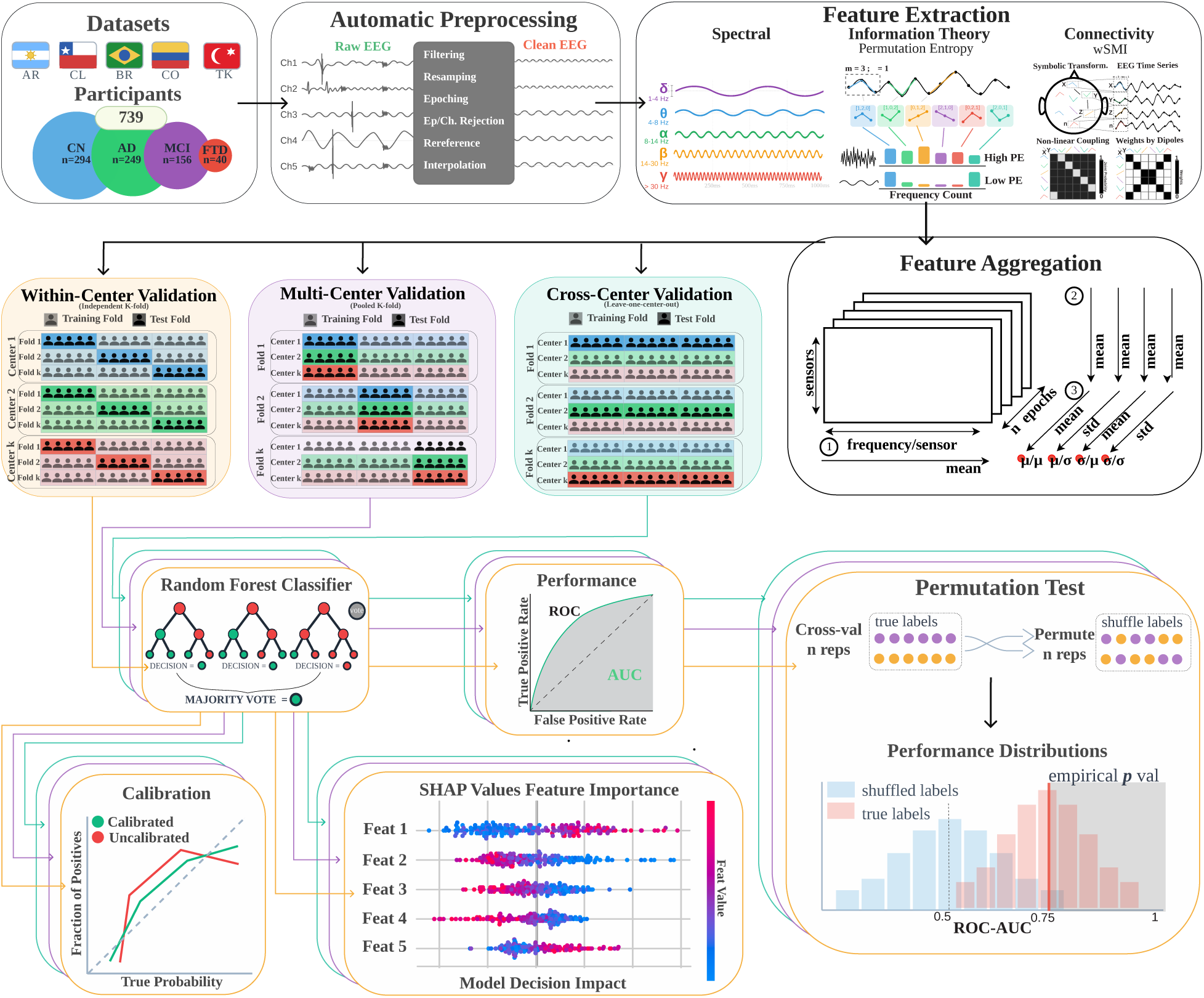
Comprehensive EEG-based machine learning pipeline for dementia classification across multiple clinical centers. The pipeline encompasses five major components: Datasets: Multi-center cohort including 739 participants across five sites (Argentina, Colombia, Brazil, Chile, Turkey) with four diagnostic groups: healthy controls (CN, n=294), Alzheimer’s disease (AD, n=249), mild cognitive impairment (MCI, n=156), and frontotemporal dementia (FTD, n=40). Automatic Preprocessing: Standardized EEG processing including filtering, downsampling, epoching, artifact rejection, re-referencing and interpolation using clean EEG protocols. Feature Extraction: Extraction of 92 neurophysiological features across three domains: spectral markers (frequency band powers), information theory measures (permutation entropy, Kolmogorov complexity), and connectivity indices (weighted symbolic mutual information) following the Sitt et al. framework. Feature Aggregation: Dual reduction approach computing spatial (mean and global field power across electrodes) and temporal (80% trimmed mean and standard deviation across epochs) statistics, yielding four feature subtypes (*µ/µ, σ/µ, µ/σ, σ/σ*) per biomarker. Machine Learning: Random Forest classification with and without probability calibration using cross-validation. Three-tier Validation: Single-center (within-site), multi-center (pooled), and cross-center (leave-one-site-out) validation strategies to assess model generalizability. Performance Evaluation: ROC-AUC analysis, reliability diagrams comparing calibrated vs. uncalibrated predictions, and statistical validation using permutation testing. Model Interpretability: SHAP values analysis for feature importance ranking and decision impact assessment. This comprehensive framework enables robust assessment of EEG biomarkers for dementia classification while addressing key translational challenges including cross-site generalizability and clinical interpretability.

## 2 Methods

### 2.1 Datasets

The current study utilized resting-state EEG data collected from five independent centers in Latin America and Europe, comprising a total of 739 participants (61% females): 294 cognitively normal (CN) individuals, 156 patients with mild cognitive impairment (MCI), 249 with Alzheimer’s disease (AD), and 40 with frontotemporal dementia (FTD), see Table 1 for the distribution between the different centers. All recordings corresponded to eyes-closed resting-state protocols. Despite differences in hardware and acquisition systems, all datasets were harmonized through a standardized preprocessing pipeline involving band-pass filtering, resampling, epoching, bad-channels and bad-epochs rejection, average reference, and bad-channel interpolation as detailed below.

**Table 1:** Demographic of participants across the five contributing centers. The table reports the number of participants (*n*), sex distribution (female:male), mean age with standard deviation (years), and mean years of education with standard deviation for each diagnostic group: cognitively normal (CN), mild cognitive impairment (MCI), Alzheimer’s disease (AD), and frontotemporal dementia (FTD). The bottom rows summarize the total distribution across all centers.

| Dataset | Variable | CN | MCI | AD | FTD |
| --- | --- | --- | --- | --- | --- |
| *BRAINLAT<sub>ar</sub>* | n | 0 | 0 | 26 | 24 |
|  | Sex (female:male) | — | — | 17:9 | 9:15 |
|  | Age (years) | — | — | 74.50 (7.50) | 69.75 (11.07) |
|  | Years of education | — | — | 9.23 (4.88) | 14.21 (4.47) |
| *BRAINLAT<sub>cl1</sub>* | n | 27 | 0 | 19 | 0 |
|  | Sex (female:male) | 15:12 | — | 10:9 | — |
|  | Age (years) | 73.93 (4.21) | — | 77.89 (9.76) | — |
|  | Years of education | 13.41 (4.50) | — | 11.32 (4.99) | — |
| *BRAINLAT<sub>cl2</sub>* | n | 30 | 0 | 38 | 16 |
|  | Sex (female:male) | 5:25 | — | 23:15 | 5:11 |
|  | Age (years) | 63.30 (10.25) | — | 69.92 (7.24) | 71.38 (5.50) |
|  | Years of education | 16.67 (3.48) | — | 13.20 (4.70) | 14.33 (3.42) |
| *IUEFM<sub>tr</sub>* | n | 98 | 100 | 97 | 0 |
|  | Sex (female:male) | 63:35 | 47:53 | 64:33 | — |
|  | Age (years) | 65.63 (8.33) | 73.62 (6.63) | 74.28 (5.12) | — |
|  | Years of education | 12.72 (4.45) | 10.34 (4.64) | 8.69 (4.44) | — |
| *USCO – UCC<sub>co</sub>* | n | 66 | 54 | 0 | 0 |
|  | Sex (female:male) | 53:12 | 46:8 | — | — |
|  | Age (years) | 60.29 (6.25) | 63.54 (7.01) | — | — |
|  | Years of education | 14.08 (5.20) | 7.43 (4.69) | — | — |
| *USP<sub>br</sub>* | n | 91 | 0 | 78 | 0 |
|  | Sex (female:male) | 56:35 | — | 50:21 | — |
|  | Age (years) | 60.63 (16.77) | — | 80.94 (8.35) | — |
|  | Years of education | 12.40 (3.47) | — | 10.65 (4.18) | — |
| **Total** | n | 312 | 154 | 258 | 40 |
|  | Sex (female:male) | 192:119 | 93:61 | 164:87 | 14:26 |
|  | Age (years) | 63.55 (11.76) | 70.08 (8.29) | 75.80 (8.00) | 70.40 (9.20) |
|  | Years of education | 13.35 (4.44) | 9.29 (4.85) | 10.16 (4.72) | 14.26 (4.05) |

Participants were recruited from centers in Argentina (*BRAINLAT_ar_*), Chile (*BRAINLAT_cl_*_1*/*2_), Turkey (Izmir University of Economics, *IUEMF_tr_*), Colombia (Universidad Surcolombiana, *USCO − UCC_co_*), and Brazil (Universidade de São Paulo, *USP_br_*). The distribution of participants across diagnostic categories and centers is presented in Table 1. Diagnostic classification followed international consensus criteria: MCI was defined according to the Petersen criteria with a Mini-Mental State Examination (MMSE) score *≥* 24; AD diagnosis fulfilled the NINCDS–ADRDA criteria for probable AD; and FTD diagnosis met the revised international consensus criteria for probable behavioral- variant FTD. All participants provided written informed consent in accordance with the Declaration of Helsinki, and all protocols were approved by the respective institutional ethics committees: INECO–San Martín de Tours (FWA00028264, Argentina); Hospital Clínico Universidad de Chile (FWA00029089, Chile); Universidad Adolfo Ibáñez (UAI) and Universidad de Santiago de Chile (USACH) for the *BRAINLAT_cl_*_2_ cohort; Universidade de São Paulo Hospital das Clínicas (FWA00001035, Brazil); Universidad de Antioquia (FWA00028864, Colombia); Hospital Universitario Hernando Moncaleano Perdomo (Bioethics and Research Committee No. 002-006, Colombia); and the Izmir University of Economics Ethics Committee (Decision 2018/05-09, Protocol 3815-GOA, Turkey).

EEG acquisition parameters for each site are summarized below:

- *BRAINLAT_ar_*: 128-channel Biosemi ActiveTwo AdBox Mk2 system, 2048 Hz sampling rate, Biosemi montage with linked mastoid reference.
- *BRAINLAT_cl_*_1_: 128-channel Biosemi ActiveTwo AdBox Mk2 system, 2048 Hz sampling rate, Biosemi montage with linked mastoid reference.
- *BRAINLAT_cl_*_2_: 128-channel Biosemi ActiveTwo system, 1024 Hz sampling rate, Biosemi montage with linked mastoid reference.
- *IUEMF_tr_*: 32-channel BrainAmp system, 500 Hz sampling rate, standard 10–20 montage.
- *USCO − UCC_co_*: 64-channel Biosemi ActiveTwo system, 1024 Hz sampling rate, Biosemi montage with linked mastoid reference.
- *USP_br_*: 21-channel EEGLAB system, 200 Hz sampling rate.

### 2.2 EEG preprocessing

To ensure cross-center comparability, all recordings were processed using a fully automated and harmonized pipeline implemented in MNE-Python (Gramfort et al., 2014) and the NICE framework (Sitt et al., 2014; Engemann et al., 2018). Non-EEG channels were removed, and signals were band-pass filtered (0.5–45 Hz) and resampled to 250 Hz to standardize the temporal resolution of frequency-dependent markers across acquisition systems. Continuous data were segmented into fixed-length epochs, followed by automated rejection of bad channels and epochs based on amplitude and variance criteria. An average reference was applied, and rejected channels were reconstructed through spherical spline interpolation to preserve montage geometry.

This standardized workflow was designed to minimize hardware- and montage-dependent variability while retaining neurophysiological information relevant for the 23 EEG markers. Data quality was comparable across sites, with mean bad channels ranging from 1.36–5.69% and bad epochs from 0.98–15.07%, and all recordings meeting predefined inclusion criteria. Signal quality was further verified using the Overall Data Quality (ODQ) index (Zhao et al., 2023), confirming no systematic differences in artifact contamination across diagnostic groups (Supplementary Methods 10.5.9). Full preprocessing specifications and quality-control criteria are provided in Supplementary Methods 10.5.

### 2.3 EEG feature extraction

Twenty-three EEG biomarkers were extracted following the framework of Sitt et al. (2014) and Engemann et al. (2018), which combines complementary descriptors of neural dynamics. Markers were organized into three families (Supplementary Table S2, Figure 1: Feature Extraction): spectral measures, indexing oscillatory slowing and power distribution; information-theoretic measures (permutation entropy and Kolmogorov complexity) capturing signal irregularity; and connectivity measures based on weighted symbolic mutual information (wSMI) to quantify nonlinear inter-regional coupling while reducing volume-conduction effects.

To obtain representations robust to montage and hardware differences, each marker was summarized along two dimensions. Across channels, we computed the mean activity and its spatial standard deviation, the latter reflecting global field power (i.e., how heterogeneous the marker is over the scalp). Across epochs, we used the 80% trimmed mean to capture typical activity and the standard deviation to quantify temporal variability. Combining these spatial and temporal summaries produced four intuitive descriptors: average activity, global field power, temporal variance, and joint spatiotemporal variability, yielding 92 feature subtypes in total.

Feature extraction was performed using identical parameters for all centers. Detailed computational definitions, frequency bands, and aggregation formulas are provided in Supplementary Methods 10.6.

### 2.4 Classification Procedure

The classification model was implemented as a scikit-learn pipeline combining a StandardScaler and a RandomForest- Classifier. While feature scaling is not required for tree-based models, its inclusion ensures consistency across potential model comparisons. The Random Forest classifier was selected due to its robustness in high-dimensional settings, as it performs implicit feature selection through random subspace sampling and reduces variance via bootstrap aggregation.

Importantly, no hyperparameter optimization was performed, and default model parameters were used throughout. This design choice was made to minimize model selection bias and reduce the risk of overfitting to specific cross-validation splits, particularly given the heterogeneity and limited sample sizes typical of multi-center clinical datasets. Rather than maximizing predictive performance, our goal was to assess the stability and generalizability of the extracted features under a minimally tuned and reproducible modeling framework.

Within this framework, three complementary validation levels were implemented to characterize performance under increasingly demanding generalization conditions (Figure 1 Within-, Multi- and Cross-Center Validation):

- **Within-center validation** assessed classification performance within each center independently to establish baseline discriminative capacity under controlled conditions. For each center containing multiple diagnostic categories, we applied Stratified by diagnostic category K-Fold cross-validation (k = 5 splits) to evaluate how well EEG markers distinguished diagnostic categories within homogeneous technical and demographic settings. This approach provided an estimate of classification performance by eliminating inter-site variability while preserving the fundamental challenge of distinguishing between diagnostic categories.
- **Multi-center validation** assessed model robustness across pooled heterogeneous datasets to evaluate performance under realistic data diversity. Data from multiple centers were combined and evaluated through Stratified by diagnostic category K-Fold cross-validation (k = 5 splits) at the patient level, allowing models to train and test on mixed populations while experiencing technical and demographic heterogeneity within each fold. This approach introduced variability from differences in EEG equipment, channel configurations, and participant demographics. The key distinction from single-center validation was that models experienced cross-site variability during training, enabling adaptation to heterogeneous conditions while maintaining exposure to all contributing centers during the training phase. Nevertheless, because the model was trained with data from the same centers on which it was evaluated, it was not blind to those technical and demographic conditions, allowing the model to learn the center’s effect. Although it learned to be robust across setups, it was not evaluated under complete generalization to unseen conditions. This validation could only be applied to diagnostic comparisons with a given contrast available from at least two different sites.
- **Cross-center generalization** evaluated model transferability to completely unseen acquisition conditions using Leave-One-Center-Out (LOCO) validation to assess external validity. In each fold, one center was entirely excluded from training and used exclusively for testing. This represented the most stringent level of generalization challenge, as models were required to transfer knowledge acquired from specific technical and demographic contexts to entirely different environments. The number of folds was constrained by the number of participating centers, with classification tasks limited to diagnostic pairs available across multiple sites. This validation strategy directly addressed the fundamental question of model generalizability across unseen heterogeneous multicentric conditions.

This hierarchical evaluation scheme allowed the study to acknowledge data-availability limitations while still providing a rigorous demonstration of generalizable performance where the evidence permitted, and a transparent delineation of scenarios in which results remained preliminary. Each validation strategy addressed progressively more stringent generalization requirements: single-center validation established fundamental biomarker discriminative capacity, multi- center validation assessed robustness to heterogeneity with adaptation opportunity, and cross-center validation evaluated true external validity transfer across independent datasets, constituting strong evidence that the 23-marker representation captures disease-related neurophysiology rather than center-specific artifacts.

### 2.5 Model evaluation, calibration, and clinical validity analyses

To assess performance stability and statistical relevance, we employed a repeated resampling and permutation framework. Specifically, 500 repetitions of stratified K-Fold-/LOCO- validation were used to estimate the distribution of performance metrics, including ROC-AUC, precision, sensitivity, and confusion matrix components (TN, FP, FN, TP), reported as *µ ± σ*. In parallel, 500 repetitions with permuted labels were used to generate null distributions of ROC-AUC under the hypothesis of no association between features and labels.

Effect size was quantified as the difference between the mean ROC-AUC obtained with true labels and the mean ROC-AUC obtained under permutation (*µ*(*AUC_real_*) *− µ*(*AUC_perm_*)). Statistical significance (*p_val_*) was estimated empirically as the proportion of permuted ROC-AUC values exceeding the mean ROC-AUC obtained with true labels, providing a non-parametric measure of how unlikely the observed performance is under the null distribution. Finally, FDR correction was applied over the the obtained *p_val_* to get the final significance *p_val_* for each validation strategy, classification contrast and center.

To evaluate the reliability of predicted probabilities, we compared uncalibrated outputs with probabilities calibrated using Platt scaling within a five-fold cross-validation framework. To obtain stable estimates of calibration performance, predicted probabilities from all 500 repetitions were pooled. This was necessary to ensure sufficient sample size for the estimation of calibration curves.

Calibration performance was quantified using mean squared error (MSE)-based metrics. Specifically, we computed *Cal_err_* as the MSE between the uncalibrated predicted probabilities and the ideal perfectly calibrated probabilities. We further quantified the benefit of calibration as *Cal*_Δ_, defined as the reduction in MSE achieved by calibration, computed as the difference between the MSE of the uncalibrated and calibrated predictions.

To assess the stability of individual-level predictions across validation regimes, we compared model-predicted probabilities obtained under within-center, multi-center, and cross-center validation. For each classification contrast, the subject-level predicted probabilities corresponding to out-of-fold test samples were averaged across repeated cross-validation iterations. Then, Spearman rank correlations were computed between pairs of validation strategies at the subject level to quantify the consistency of subject ranking across training conditions. Correlations were evaluated separately for each contrast and, when applicable, within centers. Statistical significance was assessed using the *p_val_*obtained from the Spearman rank statistic, and FDR correction using the Benjamini–Hochberg procedure was applied across all correlations to account for multiple comparisons.

To evaluate whether EEG-based predictions captured clinically meaningful variation beyond demographic effects, we assessed associations between model-predicted probabilities and neuropsychological measures after demographic adjustment. For each diagnostic contrast and validation strategy, subject-level predicted probabilities corresponding to out-of-fold test samples were averaged across cross-validation repetitions. Both predicted probabilities and cognitive scores were residualized with respect to age, sex, and years of education using linear regression. Spearman rank correlations were then computed between residualized predictions and residualized cognitive measures. Analyses were performed separately for each contrast and validation strategy, and statistical significance was assessed with FDR correction using the Benjamini–Hochberg procedure across validation strategies, cognitive measures, and contrasts.

Model interpretability was assessed using SHAP values computed in an out-of-fold manner alongside the cross-validation procedure. For each fold, models were trained on the training partition, and SHAP values were estimated on the corresponding held-out test partition using a permutation-based explainer. The background distribution was defined using the training data of each fold, ensuring that explanations were computed without information leakage.

This procedure yielded, for each validation strategy, diagnostic contrast, sample, and feature, a distribution of SHAP values across 500 repetitions. SHAP values were subsequently averaged across repetitions for each sample and classification contrast to obtain robust estimates of feature importances.

To visualize the effect of individual markers, we examined the ten features with the highest absolute mean SHAP values for each classification task, analyzing how their magnitude related to SHAP contribution—that is, whether higher or lower feature values increased the likelihood of predicting one class over another. Subsequently, the five most influential features per task were all pooled together in a radar plot to facilitate cross-task comparison.

In addition to the main analysis, we performed a sanity check procedure to rule out possible dataset and class size imbalance by downsampling all datasets and classes to the smaller available size (16) and confirmed that effects are mostly maintained (Full explanation in Supplementary Section 10.1).

For a detailed explanation regarding the model validation steps please refer to Supplementary Section 10.7.

## 3 Results

### 3.1 Robust cross, multi-center generalization

We first evaluated whether EEG-based markers support reliable diagnostic discrimination under increasingly stringent generalization conditions, spanning within-center, multi-center, and cross-center validation. *Within-center* validation assesses discrimination when training and testing are performed within the same clinical site, reflecting performance under homogeneous recording and population conditions. *Multi-center* validation evaluates robustness when models are trained and tested on pooled data from multiple centers, exposing them to heterogeneous acquisition setups and demographics during training. Finally, *cross-center* validation tests true external generalization by evaluating models on an entirely unseen center, requiring transfer to new technical and population contexts. Together, these complementary strategies allow discrimination performance, robustness to heterogeneity, and cross-site transferability to be examined separately (Section 2).

The discrimination between CN individuals and AD patients revealed robust EEG signatures across all validation strategies (Figure 2, Table 2). Performance within individual centers demonstrated strong classification across all sites (AUC 0.74–0.83). These within-center results indicated that EEG markers captured clinically relevant differences within homogeneous recording environments. Exposure to heterogeneous data during training through multi-center validation showed maintained performance across sites. This pattern suggested that diverse recording conditions during training strengthened model robustness. The most stringent cross-center validation confirmed cross-site transferability, with performance remaining largely stable across centers. Performance showed a non-significant increase between multi- and cross-center validations for *BRAINLAT_cl_*_2_, the smallest dataset, and a non-significant decrease for *IUEFM_tr_*, the largest dataset, suggesting that minor performance variations were primarily driven by differences in dataset size rather than limited model generalizability. Permutation testing confirmed that classification performance was significantly above chance at every center, indicating that predictions were not driven by site-specific artifacts. Probability estimates were also well calibrated across centers and validation strategies.

**Figure 2:**
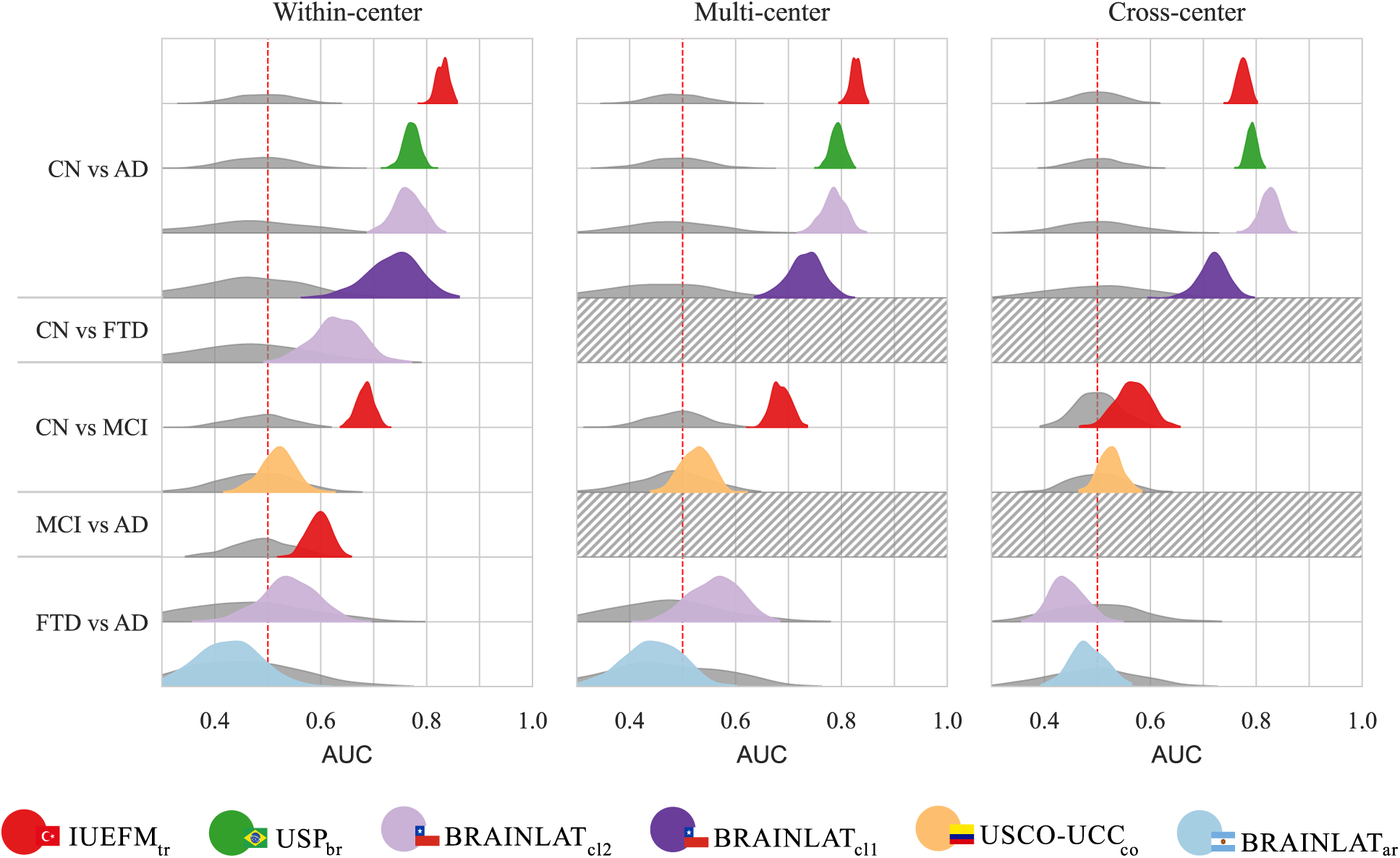
Classification performance across three validation strategies and five diagnostic comparisons. Each panel represents a different validation approach: *Within-center* (left) shows performance using Stratified by diagnostic category K-Fold cross-validation within individual centers; *Multi-center* (middle) displays results from pooled data across multiple sites using Stratified K-Fold cross-validation; *Cross-center* (right) illustrates generalization performance using Leave-One-Center-Out cross-validation, where each center was iteratively held out as an independent test set. Plots show the distribution of AUC scores across 500 repeated cross-validation iterations for each of five binary classification tasks (y-axis): CN vs AD, CN vs FTD, CN vs MCI, MCI vs AD, and FTD vs AD. Colors correspond to different acquisition centers/datasets: *BRAINLAT_ar_*(light blue), *BRAINLAT_cl_*_1_ (dark purple), *BRAINLAT_cl_*_2_ (purple), *IUEFM_tr_* (red), *USCO − UCC_co_*(orange), *USP_br_* (green). Hatched areas indicate diagnostic comparisons not available at particular centers due to absence of required diagnostic categories. The red dashed line at AUC = 0.5 marks chance-level performance.

**Table 2:**
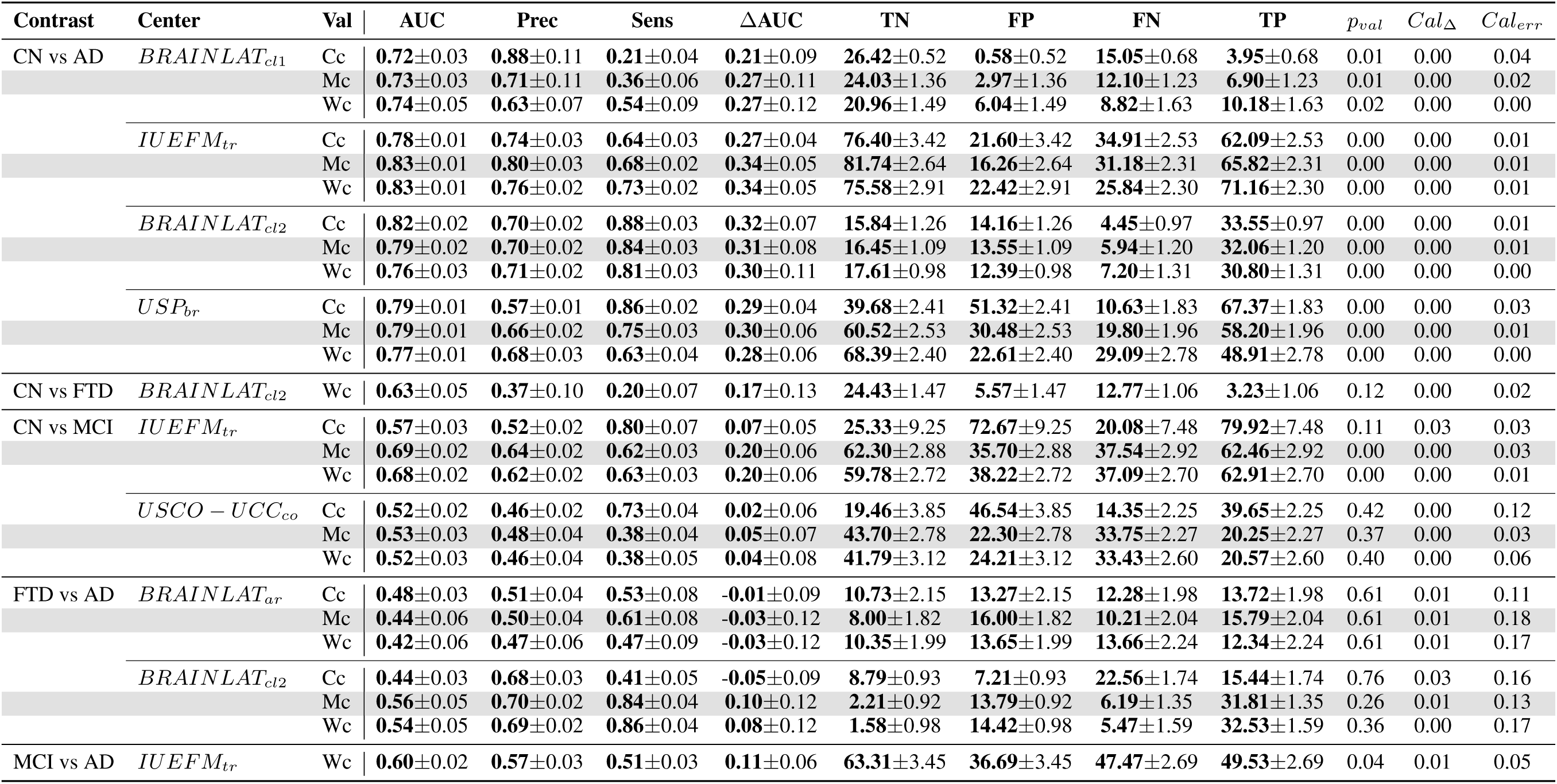
Classification performance across diagnostic contrasts, centers, and validation strategies. For each comparison, performance metrics are computed across 500 repeated runs of the full evaluation pipeline and summarized as *µ ± σ*. Reported metrics include discrimination (area under the ROC curve, AUC), precision (Prec), sensitivity (Sens), and confusion matrix components (true negatives [TN], false positives, true positives [TP]). The reported variability reflects robustness to dataset split differences and model stochasticity. In the Cross-center (Cc) validation, where train/test splits are fixed, this variability arises exclusively from model stochasticity. Effect size is quantified as the difference between the mean AUC obtained with true labels and the mean AUC obtained under permuted labels (ΔAUC). Statistical significance is assessed using a permutation-based approach, where *p_val_* corresponds to the empirical probability that the AUC obtained under permuted labels exceeds the mean AUC obtained with true labels, providing a one-sided test of performance above chance level. Calibration metrics include *Cal*_Δ_ (reduction in mean squared error after Platt scaling calibration) and *Cal_err_* (residual mean squared error between non-calibrated probabilities and observed outcomes). Three validation strategies were employed: within-center (Wc), multi-center (Mc), and cross-center (Cc).

The detection of MCI proved markedly more challenging than established dementia. Within-center analyzes showed only moderate discrimination, with substantial heterogeneity across sites, indicating that prodromal neural signatures were less consistent than those observed for CN vs AD (Figure 2, Table 2). Multi-center validation maintained the same discrimination performance, further highlighting population-specific variability in early disease expression, but with a marker signature consistent across sites. The most stringent cross-center validation further highlighted the limited generalizability of MCI detection, with performance remaining at chance level for *USCO − UCC_co_*and reaching only modest discrimination for *IUEFM_tr_*. Importantly, *USCO − UCC_co_* already exhibited near-chance performance in the within-center setting (*AUC* = 0.52), whereas training on *USCO − UCC_co_* and testing on *IUEFM_tr_* resulted in improved discrimination (*AUC* = 0.57). This finding indicates that informative patterns were present in the *USCO − UCC_co_* data, but that group separation was less pronounced within this cohort, consistent with greater heterogeneity or less distinct clinical phenotypes. Although models often achieved high sensitivity, this occurred alongside modest overall discrimination, indicating limited underlying separability rather than robust classification. Calibration quality also varied considerably between centers. Together, these findings indicate that EEG markers capture weaker and more heterogeneous neurophysiological alterations at the MCI stage.

Distinguishing between MCI and AD disease was evaluated only at *IUEFM_tr_* as we had no MCI vs AD data from the remaining centers. The within-center validation approach yielded moderate discrimination with AUC of 0.60 but statistically robust, reflecting the more challenging differentiation between advanced prodromal states and early dementia compared to CN vs AD (Figure 2, Table 2). Calibration residual error was 0.05 with improvement delta of 0.006, indicating that predicted probabilities required modest adjustment.

Distinguishing CN individuals from FTD patients was evaluated only at *BRAINLAT_cl_*_2_ as we had no CN vs FTD data from the remaining centers. Analysis within this single center achieved moderate performance with AUC of 0.63 compared with CN vs AD but statistically robust above chance level (Figure 2, Table 2). Calibration residual error was elevated at 0.02 with no significant calibration improvement, suggesting moderate deviation from optimal probability alignment. The absence of this comparison at other centers precluded multi-center and cross-center validation.

Differentiation between FTD and AD showed limited discriminative performance for *BRAINLAT_cl_*_2_ in both within- and multi-center validation strategies, while no discriminative capacity was observed for *BRAINLAT_ar_* (Figure 2, Table 2).

To further assess whether the observed effects could be influenced by imbalances in sample size across diagnostic categories and centers, we conducted an additional analysis using a repeated stratified downsampling procedure (see Supplementary Material, Section 10.1). This approach enforces identical sample sizes across all diagnostic-by-center combinations prior to model training and evaluation. The results of this analysis were consistent with the main findings (Supplementary Figure S1), indicating that the performance patterns reported above are not driven by class or center imbalance.

### 3.2 Stability of individual-level predictions across validation regimes

Beyond aggregate performance metrics, we examined whether individual-level predicted probabilities remained stable across validation regimes, thereby assessing the consistency of subject ranking under changing training conditions. For CN vs AD, predicted probabilities were highly concordant between within-, multi-, and cross-center validation strategies (Figure 3, Supplemental Table S3), indicating that participants were ranked similarly even when the model was exposed to unseen sites. This suggests that decision boundaries for established dementia relied on robust, center- independent EEG patterns. In contrast, CN vs MCI and FTD vs AD comparisons showed markedly reduced agreement when cross-center generalization was required. While within- and multi-center models produced consistent rankings, correlations involving the cross-center strategy dropped substantially, revealing more sensitivity to domain shift. These findings parallel the limited cross-center performance for these contrasts.

**Figure 3:**
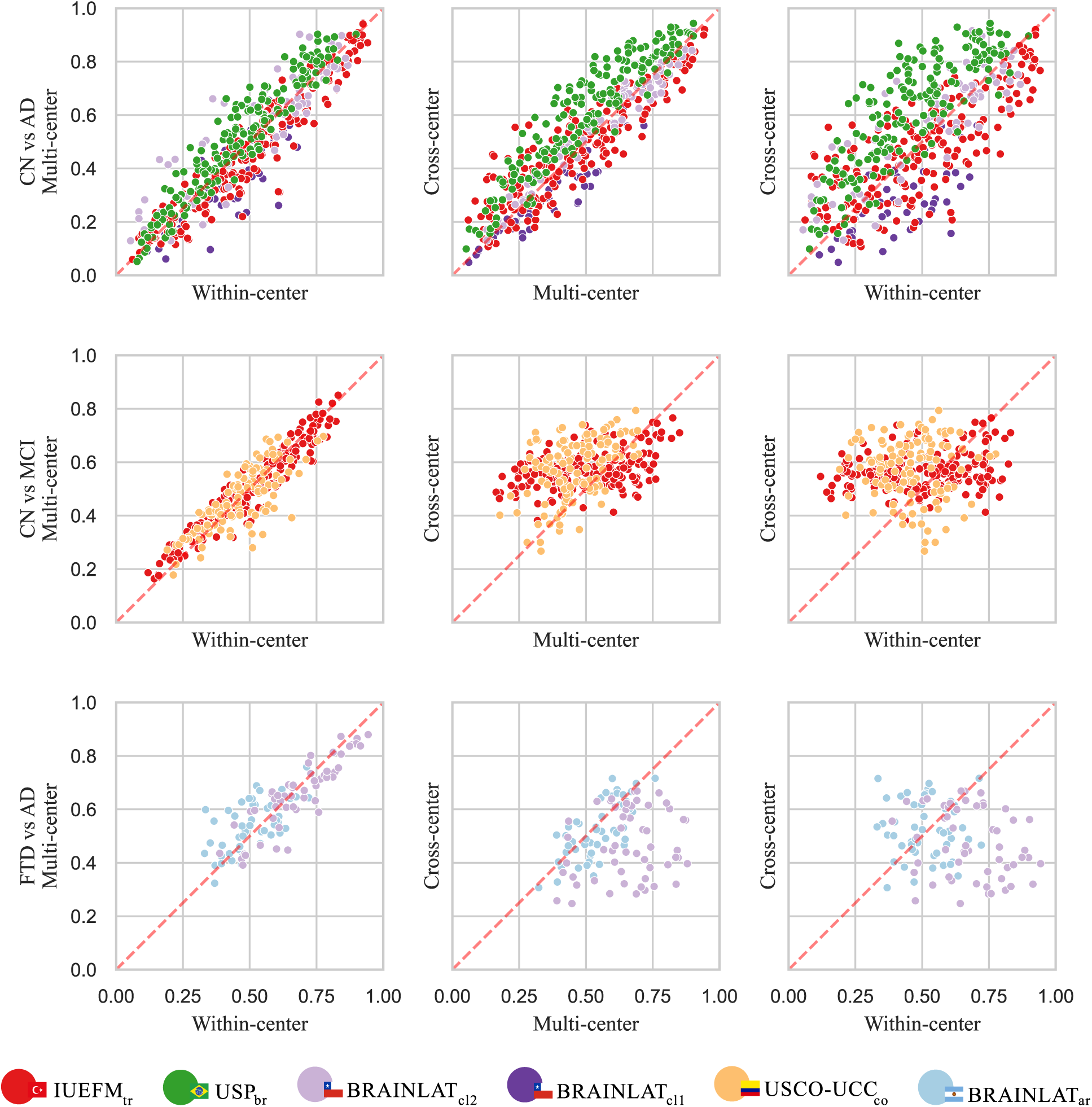
Spearman correlation matrices between paired validation strategies. Each scatter plot illustrates the relationship between predicted probabilities derived from two distinct validation strategies (columns), evaluated across different diagnostic contrasts (rows). Each point represents the mean predicted probability across repetitions for an individual participant, colored by acquisition center to visualize site-specific agreement. Spearman correlation coefficients (*ρ*) were computed for every pairwise comparison, both overall and stratified by center, to quantify the stability of decision boundaries under varying training regimes. High correlations indicate that the model maintains consistent ranking of participants regardless of the validation strategy, while low correlations suggest sensitivity to the training configuration. Complete center-specific correlation values are reported in Supplemental Table S3.

### 3.3 EEG-derived predictions align with cognitive impairment clinical tests beyond demographic effects

We first examined associations between predicted probabilities and a broad set of neuropsychological tests to characterize their overall relationship with cognitive impairment. A comprehensive analysis of these unadjusted associations, which exhibit stronger effect sizes, is provided in Supplementary Material 10.2.

Here, we focus on a more stringent analysis in which associations between predicted probabilities and cognitive measures were controlled for age, sex, and education. Also, FDR correction for multiple comparisons across cognitive measures was performed using the Benjamini–Hochberg procedure per validation strategy, classification task, and center. This approach isolates disease-related cognitive signatures from demographic effects, allowing us to test whether EEG-derived predictions capture pathology-specific patterns of impairment rather than correlations driven by demographic structure (Figure 4).

**Figure 4:**
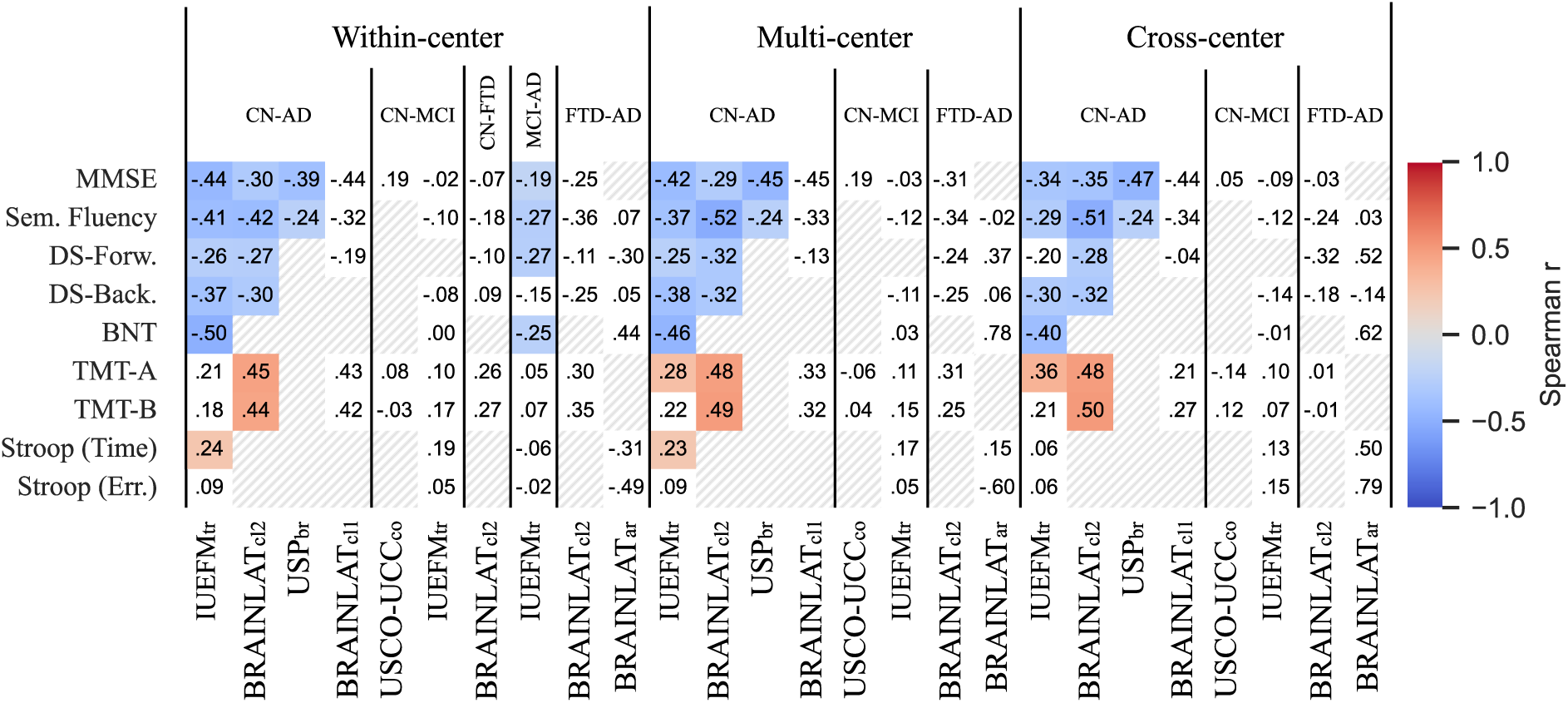
Partial Spearman correlations between model-predicted probabilities and cognitive test performance after controlling for demographic variables (age, sex, education). Each column represents a specific classification task (CN-AD, CN-MCI, CN-FTD, MCI-FTD, FTD-AD) within three validation approaches: within-center (left panel), multi- center (middle panel), and cross-center (right panel). Model predictions ranged from 0 (first class) to 1 (second class) in each binary comparison, with lower performance indicating greater impairment. Correlation coefficients are displayed within cells, with color intensity representing magnitude and direction (blue: negative, red: positive). Non colored cells indicate non significant p-values after FDR correction, while hatched cells indicate non available data. After accounting for demographic influences, negative correlations between predicted probabilities and cognitive performance (MMSE, Semantic Fluency, Digit Span, Boston Naming Test) persisted across validation schemes, demonstrating that model predictions captured disease-related cognitive decline independent of demographic factors. Positive associations with Trail Making Test remained evident, reflecting executive function deficits associated with neurodegenerative progression. The persistence of these associations after demographic adjustment suggests that EEG-derived predictions reflected genuine pathological processes rather than confounding demographic influences.

For CN vs AD, predicted AD probability showed robust and domain-consistent associations across centers and validation tiers (Figure 4, Columns 1, 6 and 9). Higher AD probability was associated with poorer global cognition (MMSE), reduced semantic fluency, lower working-memory capacity (Digit Span forward and backward), impaired confrontation naming (BNT), and slower processing speed (Stroop time) and executive functions (TMT-A and TMT-B). Most of these associations remained significant after FDR correction and were preserved in multi- and cross-center validations. These findings indicate that EEG-derived scores tracked the severity of cognitive impairment characteristic of AD, rather than reflecting site-specific or age-related effects.

In CN vs MCI, correlations did not survive FDR correction and the pattern was fragmented across centers (Figure 4, Columns 2, 7 and 10). Although several analyses revealed negative associations with memory and fluency, consistent with expected prodromal deficits, these effects were not consistently replicated. This suggests that EEG–cognition coupling in MCI is weaker and more variable across populations, reflecting the heterogeneous and transitional nature of this stage, variability in diagnostic criteria, and its partial overlap with normal aging processes.

The MCI vs AD contrast showed significant negative correlations—albeit smaller than those for CN vs AD—with MMSE, semantic fluency, Digit Span forward, and BNT (Figure 4, Column 4). Considered together, the CN vs AD, CN vs MCI, and MCI vs AD analyses delineate a coherent gradient in which CN < MCI < AD, with MCI positioned closer to CN than to AD in terms of brain–behavior coupling.

The CN vs FTD distinction, available only at *BRAINLAT_cl_*_2_ with a small sample size, did not yield robust significant correlations after FDR correction (Figure 4, Column 3), limiting interpretation of this single-center contrast. Similarly, the FTD vs AD comparison showed non-significant associations (Figure 4, Columns 5, 8 and 11). However, these findings remain exploratory given the limited FTD sample size.

Overall, the distribution of associations outlines a graded continuum consistent with cognitive proximity between conditions. The density and stability of brain–behavior links were highest for CN vs AD, decreased for CN vs MCI, and became non-significant for executive domains in FTD, paralleling the performance gradient observed in Figure 2. This convergence indicates that the EEG classifiers organize individuals according to meaningful neurocognitive structure rather than purely statistical separations.

### 3.4 Interpretable EEG markers reveal disease-specific neurophysiological signatures

SHAP values represent feature contributions to model output, with positive values pushing predictions toward the second diagnostic class and negative values toward the first class. In beeswarm plots, red points indicate high feature values and blue points indicate low values, with horizontal position showing SHAP contribution magnitude and direction (Figure 5A).

**Figure 5:**
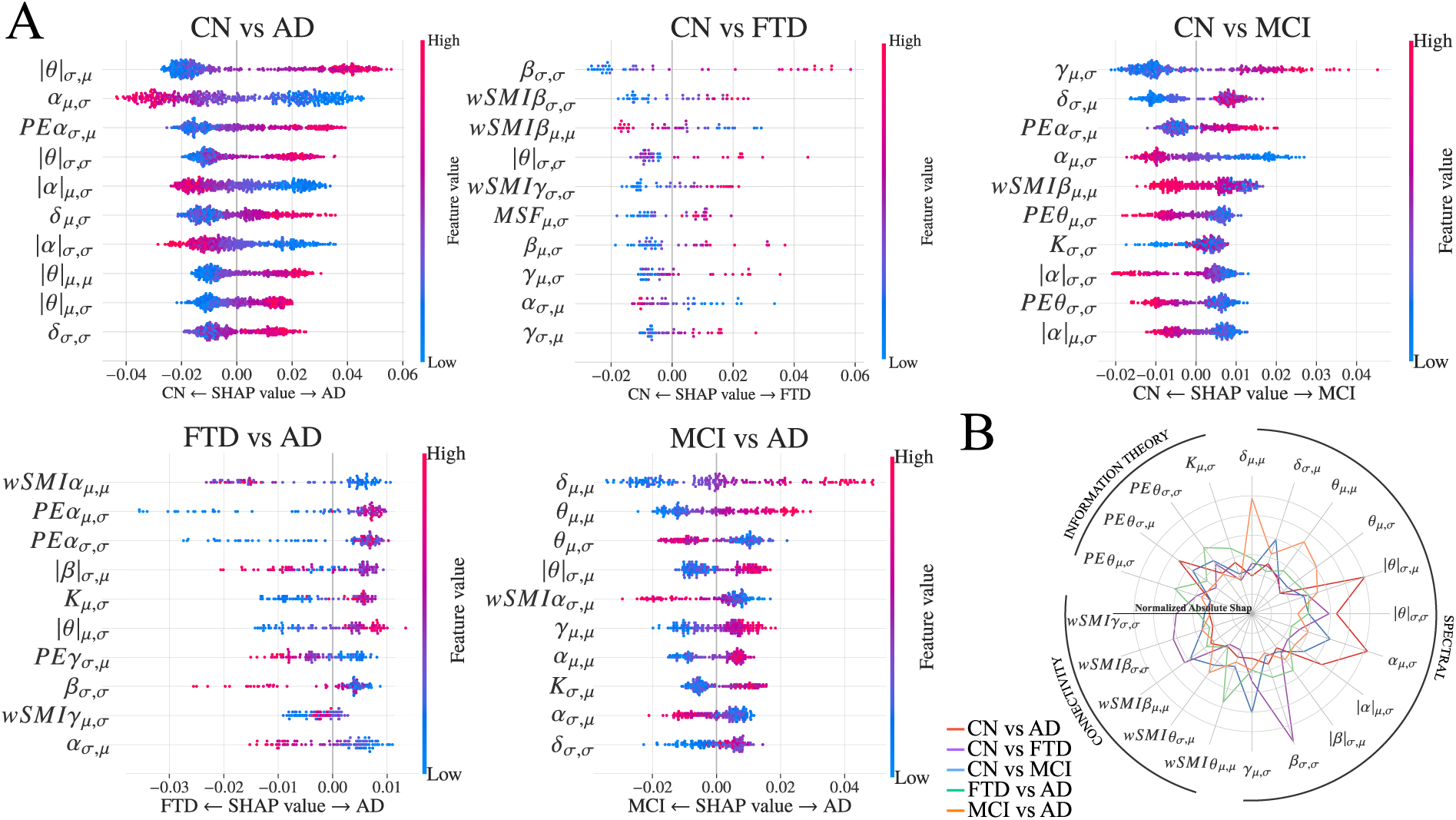
Feature importance analysis using SHAP values aggregated across all validation schemes. (A) Beeswarm plots displaying the top 10 most influential EEG features for each binary classification task. Each point represents a single prediction, with horizontal position indicating SHAP value (contribution to model output) and color reflecting feature value (blue: low, red: high). Features are ordered by mean rank of absolute SHAP values computed across all samples, pooling predictions from all validation schemes, datasets and repetitions. The notation indicates feature type and summarization method: Complexity, information theory, connectivity, and spectral markers, combined with summary statistics across channels (*µ*: mean, *σ*: standard deviation). (B) Polar plot summarizing normalized absolute SHAP values for features appearing in the top 5 of any classification task. Each axis represents a different EEG marker, with distance from the center indicating importance magnitude. Color-coded lines correspond to specific diagnostic contrasts, revealing task-specific feature utilization patterns.

The most influential features for distinguishing CN vs AD patients were characterized by two distinct, opposing neurophysiological dynamics: a global reduction in alpha-band markers and a concurrent elevation in slow-wave (theta and delta) features (Figure 5A, leftmost panel). The degradation of alpha-band activity represented a primary discriminative signature, led by the temporal variability of alpha-band power (*α_µ,σ_*). For all alpha-related markers in the top features—including spatial variability of alpha-band permutation entropy (*PEα_σ,µ_*) and normalized alpha power variability (*|α|_µ,σ_*, *|α|_σ,σ_*)—lower feature values consistently pushed predictions toward AD (positive SHAP). This convergent pattern indicated a robust reduction in both temporal and spatial signal variability within the alpha band in pathological states.

Conversely, slow-wave markers exhibited the opposite directionality, reflecting pathological cortical slowing. The spatial variability of normalized theta power (*|θ|_σ,µ_*) ranked as the most influential feature overall, matching the discriminative magnitude of alpha temporal variability. For this and all other slow-wave features among the top ten—including theta variability (*|θ|_σ,σ_*, *|θ|_µ,σ_*), average normalized theta power (*|θ|_µ,µ_*), and delta variability (*δ_µ,σ_*, *δ_σ,σ_*)—higher values consistently drove predictions toward AD. Together, these results demonstrated that the classification between CN and AD relied on a complementary interplay between elevated low-frequency activity (theta and delta) and concurrent alpha-band degradation across multiple metrics, including power, entropy, and their spatiotemporal variability (Figure 5B).

The discrimination of FTD provided a sharp counterpoint to the AD signature, relying primarily on higher-frequency alterations (Figure 5A, second panel). Reduced spatiotemporal variability of beta-band power (*β_σ,σ_*) and connectivity (*wSMIβ_σ,σ_*, *wSMIβ_µ,µ_*), alongside gamma-band markers, represented the primary discriminative features. This dominance of beta and gamma alterations contrasted clearly with the alpha-theta pattern observed in AD (Figure 5B).

Detection of prodromal decline (CN vs MCI) revealed a more heterogeneous and attenuated marker profile (Figure 5A, third panel). The discrimination relied on a mixed pattern led by reduced gamma-band variability (*γ_µ,σ_*), accompanied by moderate changes in delta power (*δ_σ,µ_*) and alpha-band permutation entropy (*PEα_σ,µ_*). Consistent with the modest classification performance, SHAP distributions for MCI showed compressed magnitude ranges and increased overlap compared to established dementia.

Finally, direct contrasts between patient groups further highlighted these distinct physiological profiles (Figure 5A, fourth and fifth panels). FTD vs AD differentiation hinged primarily on connectivity and complexity measures, specifically average alpha connectivity (*wSMIα_µ,µ_*) and permutation entropy variability (*PEα_µ,σ_*). However, SHAP distributions for this contrast showed substantial overlap, reflecting its limited discriminative capacity. Conversely, the transition from MCI to AD was characterized primarily by progressive alterations in lower-frequency markers, including elevated theta and delta power (*θ_µ,µ_*, *δ_µ,µ_*) and reduced alpha connectivity (*wSMIα_σ,µ_*).

## 4 Discussion

This study aimed to evaluate whether automated multifeature EEG-based machine learning can provide clinically meaningful, generalizable, and interpretable biomarkers for dementia across heterogeneous international cohorts. Using a fully standardized pipeline applied to five independent centers, we demonstrated that EEG classifiers achieved robust and statistically significant discrimination between CN and AD across within-center, multi-center, and leave-one-center-out validation frameworks, confirming strong external generalizability. In contrast, classification performance was progressively reduced for earlier or more heterogeneous disease stages, including MCI and differential diagnosis between dementia syndromes (AD and FTD), reflecting increasing neurophysiological overlap. Model-derived probabilistic predictions at subject-level remained stable across validation regimes and showed significant associations with cognitive impairment independent of demographic factors, supporting their clinical validity. Interpretable feature analyses revealed biologically coherent neurophysiological signatures, including alpha-band degradation and increased slow-wave activity in AD, confirming mechanistic plausibility. Together, these findings support automated EEG-based classification as a relevant approach capable of providing partially robust, clinically meaningful, and biologically interpretable biomarkers, demonstrating substantial generalizability across diverse acquisition systems and populations, and highlighting EEG as a scalable and accessible modality for dementia detection.

We captured stable neurophysiological signatures of established neurodegeneration that transfer reliably across independent centers when assessing CN vs AD, extending previous single-site and limited multi-site studies that reported promising accuracy but uncertain external validity (Akbar et al., 2025; Yuan & Zhao, 2025; Peh et al., 2021; Ohal & Mantri, 2025). This robust performance likely stems from the fact that AD represents a well-defined neurobiological entity characterized by distinct and widespread electrophysiological alterations (Babiloni et al., 2021). The preservation of classification performance under leave-one-center-out validation suggests that the extracted markers reflect disease-specific neural alterations rather than center-specific artifacts, addressing a major limitation in prior EEG machine learning research. Similarly, well calibrated, single-patient probability estimates rather than simple binary classifications, enabling clinicians to assess diagnostic confidence and guide treatment planning accordingly (Van Calster et al., 2019). Calibration results confirmed that high-performing models, particularly CN vs AD across all validation strategies, exhibited excellent baseline calibration, indicating that raw classifier outputs already approximated well-calibrated probabilities.

The reduced performance observed for MCI aligns with the known biological and clinical heterogeneity of prodromal states, where neural alterations are subtler, more variable, and partially overlap with normal aging trajectories (Saini et al., 2025; Peng et al., 2026). Such comparisons seem to be particularly vulnerable to performance degradation under stringent cross-site validation (Jiao et al., 2023; Poil et al., 2013). Similarly, the moderate performance in MCI versus AD aligns with prior reports (Farina et al., 2020), reinforcing the view of MCI not as a monolithic category but as a broad severity spectrum (Huo et al., 2025). This spectrum encompasses a range from early stages that physiologically resemble healthy aging to late-stage presentations bordering to dementia. Such heterogeneity is further compounded by diverging clinical trajectories: while a subset of these patients will eventually convert to AD, others may remain clinically stable or never progress to dementia, leading to overlapping neurophysiological signatures that complicate binary classification (Petersen et al., 2014).

Similarly, the limited discrimination between dementia subtypes (AD vs FTD) likely reflects both overlapping patho- physiological mechanisms (Ibáñez et al., 2025; Caviedes et al., 2026) and the global summary feature representation used here, which prioritizes scalability over spatial specificity. This discrepancy with prior reports (Moguilner et al., 2024; Prado, Mejía, et al., 2023) could suggest a trade-off between framework scalability and spatial resolution. By reducing scalp data to global averages, our pipeline effectively captures the widespread, randomized neural activity characteristic of AD (Babiloni et al., 2021; Ahn et al., 2025). However, this global approach likely obscures the focal, topographically restricted markers required to distinguish FTD from the posterior-dominant pathology of AD (Nishida et al., 2011; Rostamikia et al., 2024). Collectively, these findings suggest limited discrimination between dementia subtypes.

The identification of alpha slowing and increased low-frequency activity as primary discriminative features is consistent with mechanistic models of AD (Coronel-Oliveros et al., 2024), reinforcing the biological validity of the classifier. This pattern directly reflects the hallmark cortical slowing of AD (Babiloni et al., 2021; Dauwels et al., 2011), a finding further supported by the complementary elevation of delta and theta variability. In contrast, prodromal MCI revealed an intermediate profile uniquely distinguished by gamma-band variability, capturing predementia heterogeneity (Jelic et al., 2000). Discrimination of FTD provided a sharp counterpoint to AD, relying on beta and gamma markers rather than alpha slowing (Nishida et al., 2011), while FTD-AD differentiation hinged on complexity measures consistent with divergent entropic signatures (Ahn et al., 2025). Thus, these results provide a rich neurophysiological characterization of the different diseases.

The association with cognitive impairment provides clinically meaningful disease characterization (Moguilner et al., 2024; Prado, Mejía, et al., 2023) and confirms that greater cognitive impairment is linked to higher disease probability estimates (Arevalo-Rodriguez et al., 2015; Ferrante et al., 2024; Huntley & Howard, 2010). Furthermore, predictions demonstrated expected demographic patterns, with higher AD probabilities in older individuals and lower probabilities in more educated participants, consistent with education’s well-established protective role against cognitive decline (Sharp & Gatz, 2011). As the associations were preserved after controlling for age, sex, and education, EEG-derived predictions reflect pathological processes and are unlikely to be mainly driven by confounds. Collectively, these results support the view that multifeatured EEG markers can provide robust, mechanistically grounded indicators of neurodegeneration, while highlighting that prodromal and differential diagnosis remain inherently more challenging due to biological heterogeneity.

This study has several key strengths. The multi-center design spanning diverse acquisition systems, populations, and geographic regions provides unusually strong evidence of external validity compared with prior single-site or limited validation studies. The hierarchical validation strategy, including leave-one-center-out evaluation, represents a rigorous test of true generalizability rarely implemented in EEG dementia research. The use of a fully automated and standardized preprocessing and feature extraction pipeline minimizes operator-dependent variability and enhances reproducibility and scalability across clinical environments. The integration of probability calibration and cognitive association analyses demonstrates direct clinical relevance beyond binary classification, supporting the potential utility of EEG-derived predictions as quantitative biomarkers. The interpretability analysis provides mechanistic transparency by linking classifier outputs to known neurophysiological signatures of neurodegeneration. Finally, the feature representation was designed to remain compatible with heterogeneous and low-density EEG systems, enhancing feasibility for deployment in real-world and resource-limited clinical settings.

Several limitations should be considered. Sample size imbalance across diagnostic contrasts, particularly for frontotemporal dementia and certain cross-center comparisons, limited statistical power and constrained differential diagnosis evaluation. The use of spatially aggregated features, while improving robustness and scalability, likely reduced sensitivity to focal or topographically specific alterations relevant for distinguishing dementia subtypes. The conservative pipeline intentionally avoided hyperparameter tuning to minimize overfitting risk and ensure transferability to unseen populations; a trade-off that enhanced confidence in external validity at the cost of absolute performance metrics. Nevertheless, hyperparameter optimization could be applied in clinical deployment settings where maximum performance is desired. The cross-sectional design precludes direct assessment of longitudinal progression and predictive value for disease conversion, particularly for mild cognitive impairment. Diagnostic classification relied on clinical criteria without systematic or molecular biomarker confirmation (ATN Framework, Caviedes et al., 2026), which may introduce heterogeneity. Although the standardized pipeline minimizes variability, residual differences in acquisition hardware, preprocessing, and cohort characteristics may still influence performance. Finally, while the classifier demonstrated strong generalizability for established Alzheimer’s disease, performance for early detection and differential diagnosis remains limited, highlighting the need for multimodal integration and larger multicenter datasets. Future studies could leverage federated learning approaches which offer a promising avenue to address these limitations by enabling distributed model training across institutions while preserving patient privacy, achieving necessary sample sizes without data centralization (Peh et al., 2021).

In conclusion, this study demonstrates that automated multifeature EEG-based machine learning can achieve clinically meaningful and externally generalizable detection of Alzheimer’s disease across heterogeneous international cohorts while providing calibrated, biologically interpretable probability estimates linked to cognitive impairment. These findings establish EEG as a robust and scalable functional biomarker capable of supporting dementia screening across diverse clinical environments. More broadly, the results highlight the importance of rigorous cross-center validation, interpretable modeling, and clinically grounded evaluation for translating machine learning biomarkers into real-world applications. Future work integrating longitudinal data, multimodal biomarkers, and larger multicenter cohorts will further enable EEG-based precision diagnostics and scalable global dementia care.

## 5 Data Availability

The derived markers generated and analyzed in this study are publicly available in a Zenodo repository (Belloli, 2026a). These data include all computed features used for statistical analyses and model training, enabling full reproducibility of the reported results. The raw data are not publicly available due to ethical and legal restrictions related to sensitive clinical information and because they are owned by third-party institutions. Researchers interested in accessing the raw data should contact the original data providers directly and comply with their respective data access procedures and regulatory requirements.

## 6 Code Availability

All feature extraction procedures, classification pipelines, and statistical analyses required to reproduce the results reported in this study are publicly available in a Zenodo repository (Belloli, 2026b). The repository includes the full source code, a reproducible Python environment (Conda) with all required dependencies specified, and a detailed README providing step-by-step instructions for reproducing the experimental pipeline and analyses.

## Data Availability

The derived markers generated and analyzed in this study are publicly available in a Zenodo repository (https://doi.org/10.5281/zenodo.20206227). These data include all computed features used for statistical analyses and model training, enabling full reproducibility of the reported results. The raw data are not publicly available due to ethical and legal restrictions related to sensitive clinical information and because they are owned by third-party institutions. Researchers interested in accessing the raw data should contact the original data providers directly and comply with their respective data access procedures and regulatory requirements.

## Acknowledgments

AI is supported by grants from the Multi-partner consortium to expand dementia research in Latin America [ReD- Lat, supported by Fogarty International Center (FIC), National Institutes of Health, National Institutes of Aging (R01 AG057234, R01 AG075775, R01 AG21051, R01 AG083799, CARDS-NIH), Alzheimer’s Association (SG-20- 725707), Rainwater Charitable Foundation – The Bluefield project to cure FTD, and Global Brain Health Institute)], ANID/FONDECYT Regular (1250091 and 1210176 and 1220995); ANID/PIA/ANILLOS ACT210096; FONDEF ID20I10152, and ANID/FONDAP 15150012. The contents of this publication are solely the responsibility of the authors and do not represent the official views of these institutions. The funders had no role in study design, data collection and analysis, decision to publish or preparation of the manuscript.

Alfredis González Hernández and Jasmin Bonilla-Santos acknowledge Universidad Surcolombiana (USCO), Universidad Cooperativa de Colombia (UCC), and Hospital Universitario Hernando Moncaleano Perdomo for their institutional, academic, clinical, and logistical support

## 8 Authors Contributions

Research design: L.B., N.B., J.S. and A.I.; Data curation: L.B. and N.B.; EEG Preprocessing: L.B.; EEG analysis: L.B. and N.B.; Machine learning models: L.B. and N.B.; Writing—original draft: L.B., N.B., J.S. and A.I.; Writing— reviewing and editing: all authors; Project administration and funding: A.I. and J.S.; Accessed and verified data: L.B. and N.B.

## 9 Supplemental material

### 9.1 Robust cross, multi-center generalization are maintained after down-sampling for centers and class balance

**Figure S1:**
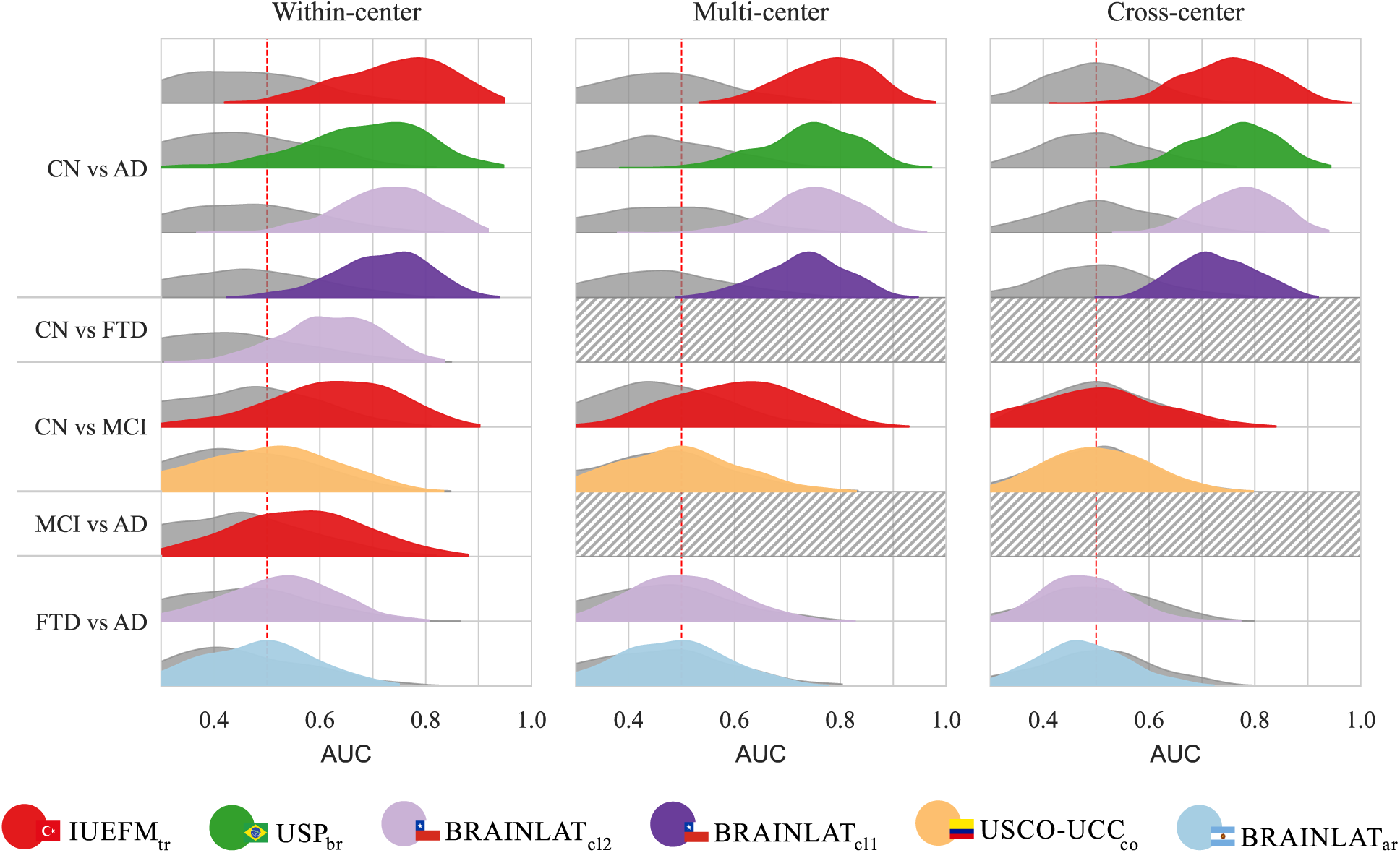
Classification performance under downsampled balanced conditions across validation strategies and diagnostic comparisons (6 samples per center and diagnostic). Each panel represents a different validation approach: *Within-center* (left) shows performance using Stratified by diagnostic category K-Fold cross-validation within individual centers; *Multi-center* (middle) displays results from pooled data across multiple sites using Stratified K-Fold cross-validation; *Cross-center* (right) illustrates generalization performance using Leave-One-Center-Out cross-validation, where each center was iteratively held out as an independent test set. Prior to model training, a repeated stratified downsampling procedure was applied to construct balanced datasets with equal sample sizes across all combinations of diagnostic category and center. Distribution plots show the AUC scores across 500 repetitions of the full pipeline (downsampling + cross-/leave-one-center-out- validations) for each binary classification tasks (y-axis): CN vs AD, CN vs FTD, CN vs MCI, MCI vs AD, and FTD vs AD. Colors correspond to different acquisition centers/datasets: *BRAINLAT_ar_* (light blue), *BRAINLAT_cl_*_1_ (dark purple), *BRAINLAT_cl_*_2_ (purple), *IUEFM_tr_* (red), *USCO − UCC_co_* (orange), and *USP_br_* (green). Hatched areas indicate diagnostic comparisons not available at particular centers due to absence of required diagnostic categories. The red dashed line at AUC = 0.5 marks chance-level performance. Overall, performance patterns closely mirror those observed in the full dataset analysis, with slightly increased variability and modest reductions in AUC consistent with reduced sample sizes.

To evaluate the extent to which the results reported in the main analysis could be influenced by imbalances in sample size across diagnostic categories and acquisition centers, we performed a repeated stratified downsampling procedure prior to model training and evaluation. For each repetition, we constructed a new balanced dataset by randomly sampling, without replacement, an equal number of observations from each combination of diagnostic category and center. The number of samples per subgroup was fixed at 16 across all centers and classes and determined by the smallest available subgroup to ensure feasibility. This procedure resulted in datasets with identical class and center distributions across repetitions, thereby removing potential confounding effects arising from unequal sample sizes.

For each resampled dataset, the same validation strategies described in the main analysis (See section 2.4) were applied using identical model configurations and evaluation pipelines.

The results of this control analysis are presented in Supplementary Figure S1. Overall, the performance patterns were highly consistent with those observed in the main analysis without downsampling. In particular, the relative differences across centers, diagnostic comparisons, and validation strategies were preserved, supporting the robustness of the main findings.

Two expected differences emerged as a direct consequence of the downsampling procedure. First, variability across repetitions increased relative to the main analysis. This reflects the fact that each repetition is performed on a different subset of participants, whereas in the original analysis all repetitions are conducted on the same dataset, with variability arising solely from differences in cross-validation splits and model stochasticity. Second, a modest decrease in performance was observed across conditions. This reduction is expected given the smaller number of training samples available in each repetition, which limits model capacity. Importantly, this decrease was relatively small and not proportional to the magnitude of downsampling, despite some datasets being substantially reduced in size (e.g., from 78,91 or 100, to 16 samples), indicating that the learned patterns remain robust even under constrained sample sizes.

**Figure S2:**
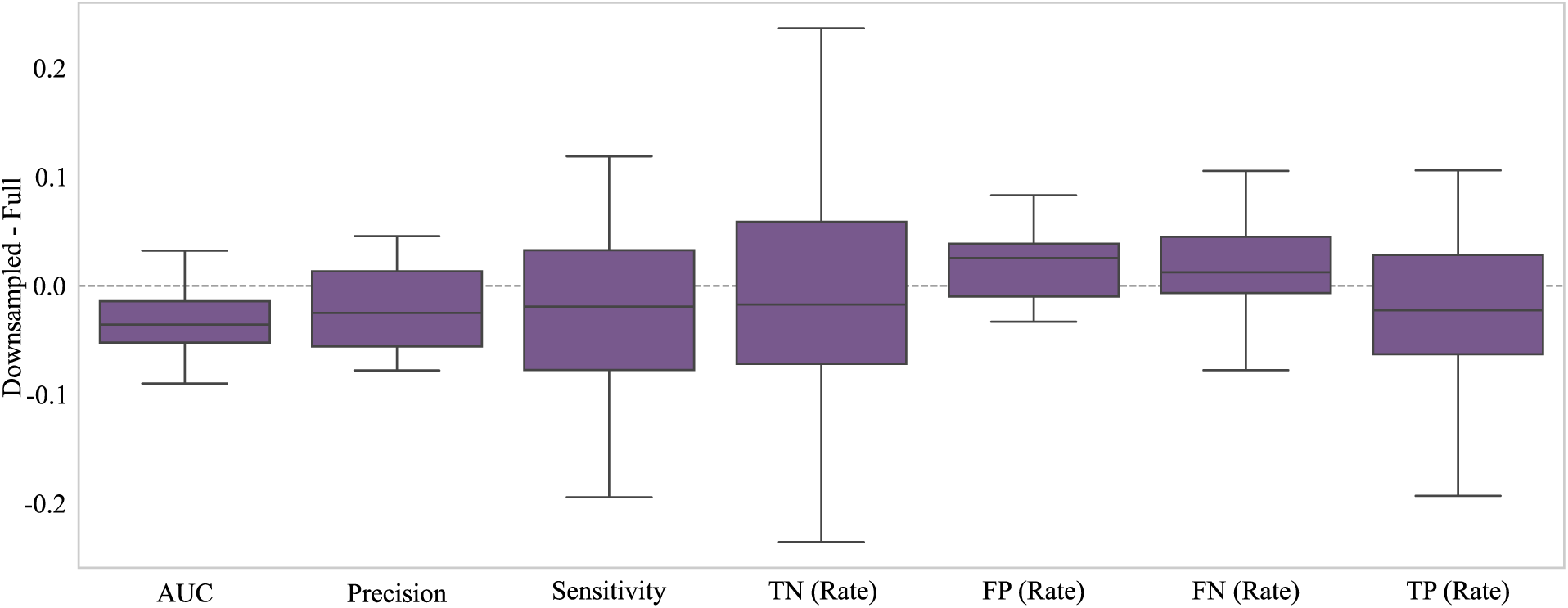
Comparison of performance metric distributions between the original and downsampled analyses across all centers, diagnostic contrasts, and validation strategies. For each metric (AUC, precision, sensitivity, and confusion matrix rates), distributions are shown for the full dataset and the downsampled dataset.

To further quantify the agreement between the original and downsampled analyses, we examined the paired differences in performance metrics across all centers, diagnostic comparisons, and validation strategies (Supplementary Figure S2). For each metric, we computed the difference between the downsampled and original results, such that values centered around zero indicate minimal impact of downsampling.

Across all evaluated metrics, the distributions of paired differences were just a small apart from zero (no differences). While some variability was observed—particularly for sensitivity and true negative rates—the overall magnitude of these deviations remained small. Importantly, no systematic degradation in performance was evident across metrics.

Taken together, the near-zero centering of the distributions and the limited spread of differences provide converging evidence that downsampling does not meaningfully alter model performance. These findings indicate that the performance patterns observed in the main analysis are not driven by imbalances in sample size across centers or diagnostic groups, but instead reflect robust and consistent signals that persist under controlled, balanced sampling conditions.

### 9.2 EEG-derived predictions align with cognitive impairment clinical tests

To evaluate whether model-predicted probabilities captured clinically meaningful patterns of cognitive and demographic variation, Spearman correlations were computed between predicted probabilities and available demographic and neuropsychological variables across all validation strategies and diagnostic comparisons. Model predictions ranged from 0 (indicating the first diagnostic class) to 1 (indicating the second class) in each binary comparison. Cognitive test scores were inverted when necessary to ensure consistent directionality, such that lower scores uniformly reflected greater impairment. Statistical significance was determined with FDR correction using the Benjamini–Hochberg procedure to account for multiple comparisons per validation strategy, classification contrast, and center.

#### 9.2.1 CN vs AD Classification

Demographic variables showed selective associations with predicted AD probabilities across validation strategies (Figure S3, leftmost columns). Age demonstrated positive correlations at specific centers, with values ranging from 0.29 to 0.34 in within-center validation, 0.34 to 0.37 in multi-center validation, and 0.32 to 0.35 in cross-center validation, indicating that older individuals at these sites received higher AD probabilities independent of diagnostic group. Education showed minimal significant associations after FDR correction, with most correlations non-significant, suggesting that the primary discriminative features captured by EEG markers operated largely independently of educational attainment.

Cognitive performance revealed substantially stronger associations with predicted probabilities across all validation strategies. Global cognitive screening through Mini-Mental State Examination showed robust negative correlations, ranging from -0.25 to -0.54 within individual centers, -0.33 to -0.55 with multi-center training, and -0.30 to -0.49 when tested on unseen sites (Figure S3). These patterns indicated that individuals with lower cognitive function consistently received higher AD probabilities. Semantic fluency paralleled this pattern with significant negative correlations across validation strategies (*r_wc_* = -0.28 to -0.51, *r_mc_* = -0.34 to -0.50, *r_cc_* = -0.22 to -0.50), suggesting that verbal fluency deficits characteristic of Alzheimer’s disease corresponded reliably with EEG-based probability estimates.

Working memory capacity demonstrated differential sensitivity depending on task demands. Backward digit span showed stronger associations (*r_wc_* = -0.25 to -0.46, *r_mc_* = -0.38 to -0.48, *r_cc_* = -0.39 to -0.41) than forward span (*r_wc_* = -0.20 to -0.21, *r_mc_* = -0.21 to -0.32, *r_cc_* = -0.13 to -0.33), with this differential pattern suggesting particular sensitivity to executive control aspects of working memory. Confrontation naming through Boston Naming Test revealed strong negative correlations at *IUEFM_tr_*, particularly evident in within-center (*r_wc_* = -0.58), multi-center (*r_mc_* = -0.56), and cross-center (*r_cc_*= -0.48) validation, indicating that semantic memory impairment aligned consistently with EEG-derived probability estimates.

Processing speed and executive function showed positive relationships with disease probabilities, as completion times increase with cognitive decline (Figure S3). Trail Making Test A demonstrated significant associations (*r_wc_* = 0.31-0.38, *r_mc_* = 0.34-0.42, *r_cc_* = 0.19-0.42), while Trail Making Test B showed similar patterns (*r_wc_* = 0.25-0.28, *r_mc_* = 0.35-0.44, *r_cc_* = 0.26-0.46). These associations persisted across validation strategies, indicating that processing speed deficits corresponded reliably with predicted AD probabilities.

#### 9.2.2 CN vs MCI Classification

The distinction between cognitively normal individuals and those with mild cognitive impairment revealed more variable and generally weaker cognitive-prediction associations (Figure S3, second columns). Demographic variables demonstrated minimal significant associations after FDR correction, with age showing small positive correlations at *IUEFM_tr_* (*r_wc_* = 0.25, *r_mc_* = 0.22) and education remaining non-significant across centers. Mini-Mental State Examination showed variable negative correlations across centers and validation strategies, ranging from -0.25 to -0.42 within centers, -0.33 to -0.49 with multi-center training, and -0.30 to -0.49 when tested on unseen sites.

Semantic fluency demonstrated negative correlations primarily at *IUEFM_tr_* across validation strategies (*r_wc_* = -0.28, *r_mc_* = -0.48, *r_cc_* = -0.50), while working memory measures showed selective associations with digit span backward (*r_wc_* = -0.25, *r_mc_* = -0.38, *r_cc_* = -0.39) at *IUEFM_tr_*. Processing speed measures revealed patterns concentrated at specific centers (Figure S3). Trail Making Test A (*r_wc_* = 0.38, *r_mc_* = 0.42, *r_cc_*= 0.42) and Test B (*r_wc_* = 0.25, *r_mc_* = 0.44, *r_cc_* = 0.46) showed significant positive correlations at *IUEFM_tr_* across validation strategies, indicating that processing speed and executive function deficits showed correspondence with EEG-based predictions at specific centers.

#### 9.2.3 Other Diagnostic Comparisons

The CN vs FTD distinction, available only at *BRAINLAT_cl_*_2_, demonstrated moderate associations with age (*r_wc_* = 0.29), Mini-Mental State Examination (*r_wc_* = 0.26), and processing speed measures (Figure S3, third column of within-center panel). Interpretation remained limited by single-center availability. The MCI vs AD comparison revealed significant associations with Mini-Mental State Examination (*r_wc_* = -0.15), semantic fluency (*r_wc_* = -0.20), and Trail Making Test performance (*r_wc_* = 0.22-0.27), aligning with the moderate discriminative performance observed for this diagnostic comparison.

The FTD vs AD differentiation revealed minimal significant associations with demographic or cognitive measures after FDR correction across validation strategies (rightmost columns, Figure S3). Most cognitive measures including Mini-Mental State Examination, semantic fluency, and digit span showed predominantly non-significant correlations. Trail Making Test demonstrated scattered significant associations at *BRAINLAT_ar_* (*r_wc_* = 0.40-0.49, *r_mc_* = 0.23-0.36, *r_cc_* = 0.80 for Test B), though patterns were inconsistent across sites. This sparse pattern corresponded with the limited discriminative capacity observed for this diagnostic comparison.

**Figure S3:**
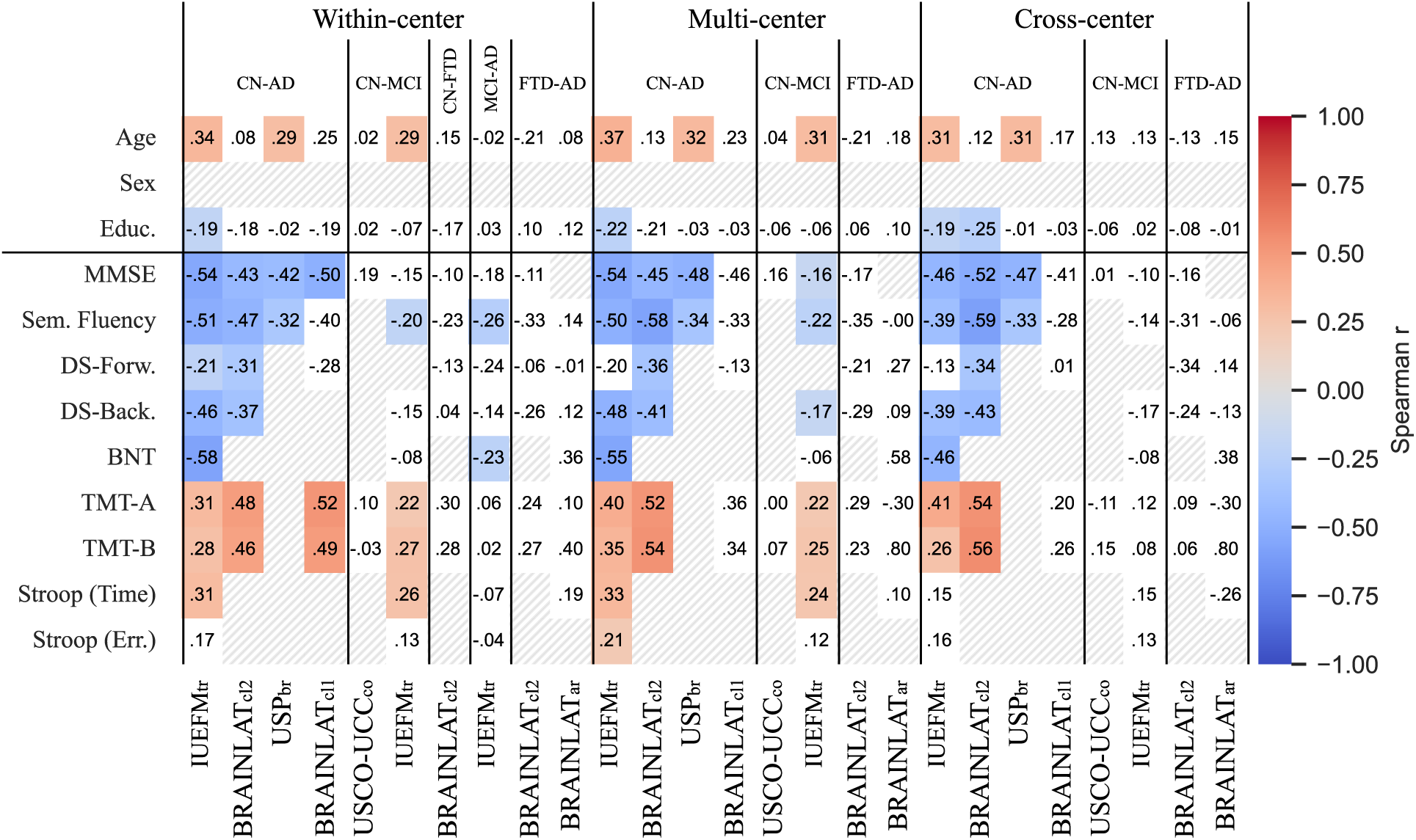
Spearman correlations between model-predicted probabilities and demographic/cognitive measures across validation schemes. Each column represents a specific classification task (CN-AD, CN-MCI, CN-FTD, MCI-FTD, FTD-AD) within three validation approaches: within-center (left panel), multi-center (middle panel), and cross-center (right panel). Model predictions ranged from 0 (first class) to 1 (second class) in each binary comparison. Cognitive test scores were inverted when necessary to ensure consistent directionality, with lower performance indicating greater impairment. Correlation coefficients are displayed within cells, with color intensity representing magnitude and direction (blue: negative, red: positive). Hatched cells indicate non-significant correlations after false discovery rate correction.

### 9.3 Feature importance stability across cross-validation repetitions

To assess whether the EEG features identified as most influential by SHAP analysis were consistently selected across repeated cross-validation runs, rather than being artefacts of particular train/test splits, we computed the Jaccard similarity of top-10 feature sets across all pairwise repetition pairs. For each repetition and each diagnostic contrast, the top 10 features were identified by ranking features according to their mean absolute SHAP value on the held-out test fold. The Jaccard index between each pair of repetitions was then computed as the cardinality of the intersection of the two feature sets divided by the cardinality of their union. This procedure was applied independently for each diagnostic contrast and validation strategy, yielding a distribution of pairwise Jaccard values whose mean and standard deviation quantify the stability and reproducibility of the identified feature signatures.

Results are presented in Supplementary material Table S1 and visualised in Supplementary material Figure S4. Feature importance stability mirrored the classification performance gradient described in the main analysis. For CN vs AD, Jaccard similarity was high across validations (0.77 to 0.89), indicating that the same subset of features (primarily alpha-band degradation and slow-wave increases) was reliably recovered regardless of the training configuration. This consistency supports the interpretation that the discriminative signature for established Alzheimer’s disease is robust and not sensitive to dataset composition, reinforcing the biological plausibility of the identified biomarkers.

For CN vs MCI, feature stability was reduced across all validation strategies but maintained considerable stability (0.38 to 0.66). This stability range is consistent with the heterogeneous and attenuated neurophysiological alterations at the prodromal stage and echoes the modest and variable classification performance for this contrast. The particularly low mean Jaccard value for the Multi-center case suggests that the feature landscape shifts substantially when models are trained on pooled heterogeneous data, likely because different centers capture different facets of early neurodegeneration in their MCI cohorts. A similar pattern of reduced stability was observed for FTD vs AD (0.33 to 0.53) and MCI vs AD (0.50), consistent with the limited and heterogeneous discriminative capacity observed for these contrasts in the main analysis. The CN vs FTD contrast, available only within *BRAINLAT_cl_*_2_, showed intermediate stability (0.61).

Taken together, these findings demonstrate that SHAP-identified biomarkers are reproducible in proportion to the discriminability of the underlying contrast. High feature stability for CN vs AD across validation regimes provides additional evidence that the alpha-theta signature reported in the main analysis is a genuine, center-independent neurophysiological marker of established Alzheimer’s disease. The graded reduction in stability for MCI-related and FTD contrasts further underscores the inherently more variable neural phenotypes at these diagnostic boundaries, and highlights the need for larger multicentric samples and more targeted feature representations to achieve reliable identification of prodromal and differential dementia biomarkers.

**Figure S4:**
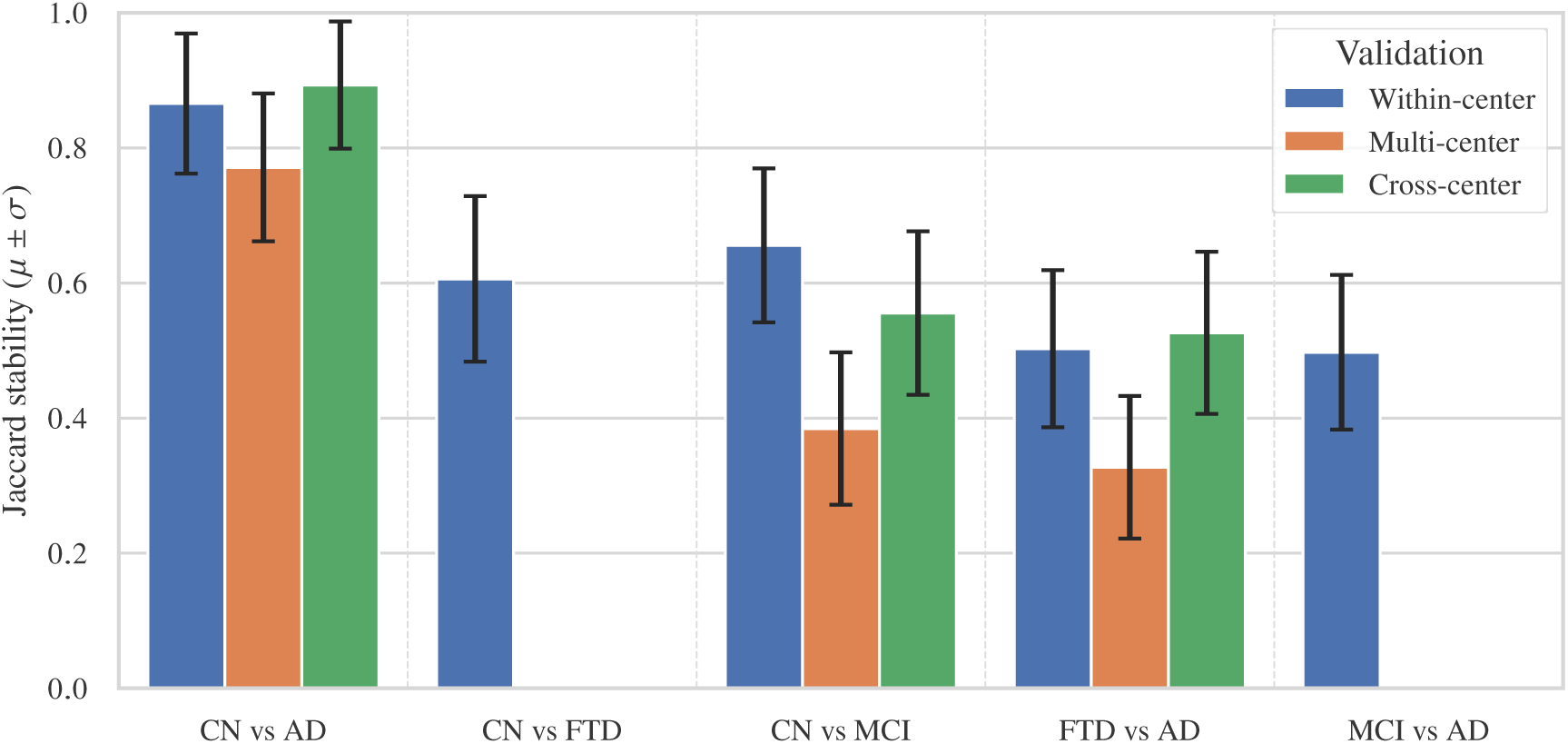
Jaccard similarity of top-10 feature sets (ranked by mean absolute SHAP value on the held-out test fold) across repeated cross-validation runs, shown for each diagnostic contrast and validation strategy (within-center, multi- center, and cross-center). For each repetition pair, the Jaccard index was computed as the size of the intersection divided by the size of the union of the two top-10 features, ranging from 0 (no overlap) to 1 (identical sets). Bars show the mean Jaccard similarity across all pairwise repetition comparisons; error bars indicate standard deviation from the mean. Absent bars indicate validation strategies not available for a given contrast.

### 9.4 Feature importance under downsampled balanced conditions

To verify that the neurophysiological signatures identified by SHAP analysis in the main analysis were not driven by imbalances in sample size across diagnostic categories and acquisition centers, we replicated the feature importance analysis using the same downsampled balanced datasets described in Section 10.1, in which an equal number of observations (n = 16) was drawn per diagnostic and center combination prior to model training.

The resulting SHAP profiles are presented in Supplementary material Figure S5. Feature importance patterns were highly consistent with those reported in the main analysis (Figure 5). For CN vs AD, the two opposing neurophysiological dynamics remained the dominant discriminative signature: alpha-band degradation — led by the temporal variability of alpha power (*α_µ,σ_*) and alpha permutation entropy (*PEα_σ,µ_*) — and concurrent elevation of slow-wave theta and delta features, with *|θ|_σ,µ_* retaining its rank as the single most influential feature. The directionality of all top features was unchanged relative to the full-sample analysis. For CN vs FTD, beta-band power and beta-band connectivity (*wSMIβ*) continued to dominate, with median spectral frequency (*MSF_µ,σ_*) appearing somewhat more prominently than in the full-sample analysis, consistent with the spectral slowing expected in this contrast. For CN vs MCI, the heterogeneous and attenuated profile was preserved, with gamma-band variability (*γ_µ,σ_*) again leading, accompanied by delta and alpha entropy features; beta-band connectivity features appeared with slightly greater weight under balanced sampling, suggesting a modest contribution that may have been partially obscured by class imbalance in the full dataset. For FTD vs AD and MCI vs AD, the connectivity- and complexity-driven profiles described in the main text were likewise preserved, with delta and theta features maintaining their leading role in the MCI-to-AD transition.

**Table S1:**
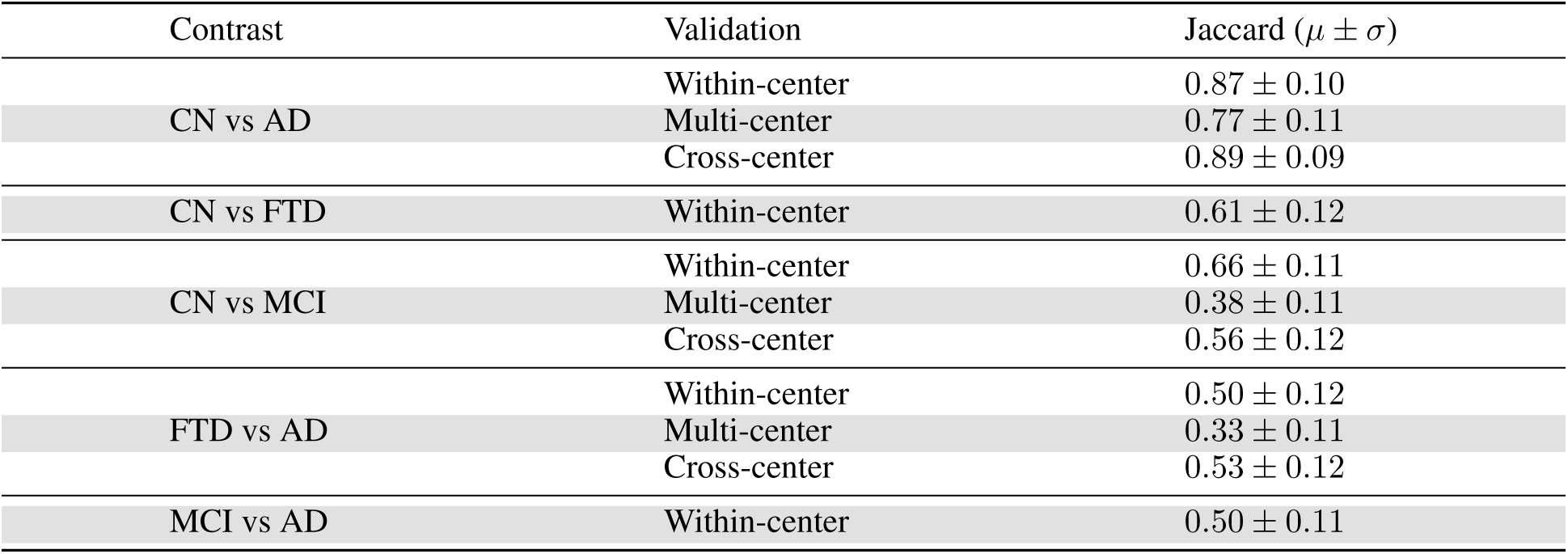
Jaccard similarity of top-10 feature sets (ranked by mean absolute SHAP value on the held-out test fold) across repeated cross-validation runs, shown for each diagnostic contrast and validation strategy (within-center, multi-center, and cross-center). For each repetition pair, the Jaccard index was computed as the size of the intersection divided by the size of the union of the two top-10 features, ranging from 0 (no overlap) to 1 (identical sets). Table shows the mean and standard deviation of the Jaccard similarity across all pairwise repetition comparisons.

Across all contrasts, absolute SHAP magnitudes were modestly reduced relative to the full-sample analysis, consistent with the smaller training set sizes available under downsampling. Taken together, these results confirm that the disease-specific neurophysiological signatures identified in the main analysis are not artefacts of class or center imbalance, but reflect genuine and robust patterns in the EEG data.

### 9.5 Extended EEG preprocessing

All preprocessing was performed offline using a fully automated pipeline implemented in **MNE-Python** and the **NICE** framework, following procedures established for multi-center clinical EEG harmonization Sitt et al., 2014; Engemann et al., 2018. The pipeline was applied identically to all datasets without site-specific tuning.

#### 9.5.1 Channel selection

Non-scalp and non-EEG channels were excluded, including electrooculographic (EOG), electromyographic (EMG), external reference, and system status channels. Only scalp EEG channels were retained for further processing.

#### 9.5.2 Filtering

Signals were band-pass filtered between **0.5–45 Hz** using zero-phase IIR Butterworth filters. Implementation consisted of a **4th-order high-pass filter at 0.5 Hz** followed by an **8th-order low-pass filter at 45 Hz**. Both filters were applied with forward–backward (filtfilt) correction to eliminate phase distortion.

#### 9.5.3 Resampling and trimming

All recordings were **resampled to 250 Hz** to ensure comparability of frequency-dependent markers across acquisition systems. The initial **0.8 s of each recording** were removed to exclude start-up transients.

#### 9.5.4 Epoching

Continuous data were segmented into **0.8-s epochs** with **50% overlap**, providing stable estimation for spectral and information-theoretic markers.

#### 9.5.5 Adaptive artifact rejection

Artifact detection followed a four-step procedure, applied sequentially to channels and epochs Engemann et al., 2018. Each step operated on peak-to-peak amplitude or variance statistics computed directly from the epoched data.

**Figure S5:**
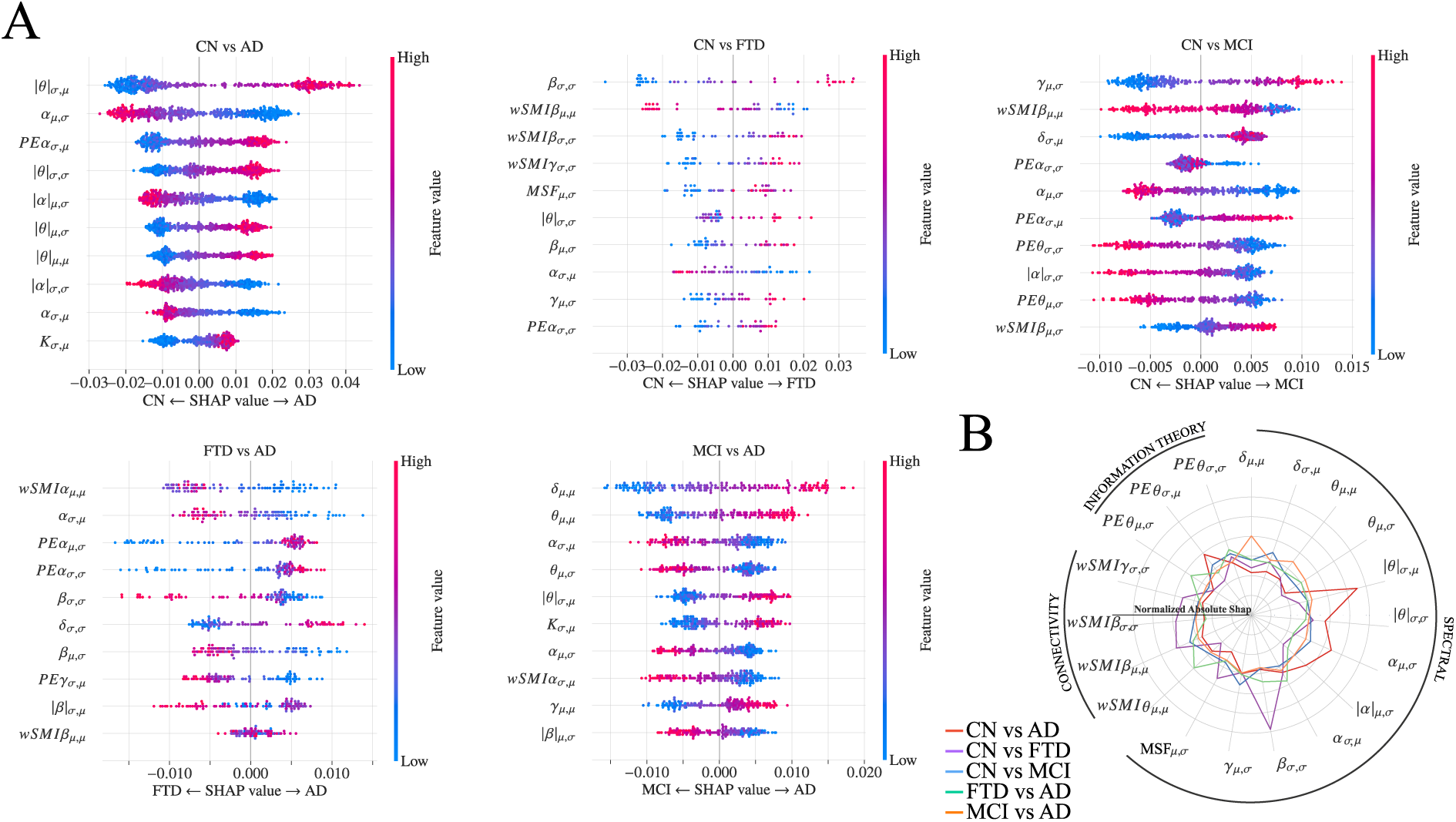
Feature importance analysis using SHAP values under downsampled balanced conditions (n = 16 per diagnostic and center). Layout and notation follow Figure 5 of the main text. (A) Beeswarm plots showing the top-10 most influential EEG features for each binary classification task, with SHAP values averaged across all validation schemes and 500 repeated downsampling-plus-cross-validation iterations. Each point represents a single prediction; horizontal position indicates the SHAP contribution magnitude and direction, and color reflects the feature value (blue: low; red: high). Features are ordered by mean rank of absolute SHAP values. (B) Polar plot summarising normalised absolute SHAP values for features appearing in the top 5 of any classification task, with color-coded lines corresponding to each diagnostic contrast. Feature importance patterns are highly consistent with the full-sample analysis (Figure 5), confirming that the identified neurophysiological signatures are robust to class and center imbalance. Modest reductions in absolute SHAP magnitudes reflect the smaller training set sizes inherent to the downsampling procedure.

1. **Channel rejection by peak-to-peak range**. For each channel, the range (maximum minus minimum) was computed within every epoch. Channels were marked as bad when the number of epochs with a range exceeding **100 µV** was greater than **50% of all epochs**. This criterion identifies sensors with recurrent large-amplitude fluctuations across time.
2. **Channel rejection by variance outliers**. Channel variances were estimated across all samples, and an iterative Z-score procedure was applied using an outlier detection mechanism. Channels whose variance exceeded a threshold of **4 standard deviations** from the median were flagged as bad, with up to **4 iterations** to remove local outliers before recomputation.
3. **Epoch rejection by channel contamination**. For each epoch, the proportion of channels whose peak-to-peak range exceeded **100 µV** was calculated. Epochs were rejected when more than **10% of channels** violated this threshold, ensuring that transient artifacts affecting multiple sensors were removed.
4. **Channel rejection by high-frequency variance**. To capture residual muscle or line-noise contamination, data were high-pass filtered at **25 Hz** (4th-order Butterworth). Channels with abnormally large standard deviation in this high-frequency band were detected using the same iterative Z-score procedure (**4 SD**, **max 4 iterations**).

The procedure was terminated if all channels or all epochs were flagged as bad at any intermediate step. The final set of bad channels was the union of those detected across the three channel-level criteria.

#### 9.5.6 Inclusion criteria

Recordings were retained only if at least **200 artifact-free epochs** remained and more than **70% of original EEG channels** were usable after rejection. All recordings passed inclusion criteria.

#### 9.5.7 Re-referencing and interpolation

Data were re-referenced to the **average reference** computed from all non-rejected channels. Bad channels were reconstructed using **spherical spline interpolation** to preserve the original montage geometry.

#### 9.5.8 Implementation notes

All steps were executed automatically with fixed parameters across centers to avoid site-specific optimization. The pipeline was applied prior to feature extraction without access to diagnostic labels, ensuring independence between preprocessing and subsequent machine-learning analyses.

#### 9.5.9 Overall Data Quality (ODQ) analysis

To verify that the preprocessed EEG recordings did not exhibit systematic differences in signal quality across diagnostic groups, which could confound downstream feature extraction, we computed the Overall Data Quality (ODQ) index for each EEG recording following the methodology proposed by (Zhao et al., 2023). ODQ quantifies the proportion of artifact-free signal segments and is derived from four complementary detection methods applied to the epoched data (0.8 seconds). The four methods flag windows containing: (i) constant or flat signals (ONS; detected via standard deviation and median absolute deviation thresholds below 10*^−^*^10^); (ii) high-amplitude transients (OHA; detected via an absolute amplitude threshold of 150 µV and a z-scored robust standard deviation threshold); (iii) high-frequency noise (OFN; detected via the noise-to-signal ratio relative to the power-line frequency cutoff, with both absolute and z-scored thresholds); and (iv) low inter-channel correlation (OLC; detected when the maximum Pearson correlation between a channel and all others fell below 0.6). A window was classified as good quality only when it passed all four criteria, and ODQ was expressed as the percentage of good windows over the total number of windows.

To test whether ODQ differed systematically across diagnostic categories, pairwise comparisons between all condition pairs (CN, MCI, AD, FTD) were performed using two-sided Mann-Whitney U tests, both globally across all datasets pooled and separately within each of the six participating centers.

No significant differences in ODQ were found between any pair of diagnostic conditions, either in the pooled analysis (Figure S6 or within individual datasets (Figures S7). These results confirm that the preprocessed recordings are balanced in signal quality across diagnostic groups and that observed differences in EEG features are unlikely to be attributable to differential artifact contamination.

#### 9.6 Extended EEG feature extraction

EEG biomarkers were computed using the NICE Python library following the framework of Sitt et al. (2014) and Engemann et al. (2018). Feature extraction was applied identically to all centers and exclusively on preprocessed data, without access to diagnostic labels.

#### 9.6.1 Marker families

##### Spectral markers

Power spectral density was estimated for each channel and epoch using Welch’s method. Features were computed in canonical frequency bands: *δ* (1.25–4 Hz), *θ* (4–8 Hz), *α* (8–13 Hz), *β* (13–30 Hz), and *γ* (30–45 Hz). Derived descriptors included absolute and relative band power, median spectral frequency, spectral edge (90/95%), and spectral entropy. It’s worth noticing that the *δ* band is calculated from 1.25Hz because epochs with 0.8 seconds length make it not possible to calculate lower frequencies.

##### Information-theoretic markers

Signal irregularity was quantified using permutation entropy (PE) with embedding dimension *m* = 3 and band-specific delays *τ* matched to the center frequency of each band. Kolmogorov complexity (K) was estimated to capture algorithmic compressibility of the time series.

##### Connectivity markers

Inter-electrodes coupling was assessed using weighted symbolic mutual information (wSMI). After symbolic discretization of the signals, mutual information was computed for all channel pairs, providing a nonlinear connectivity measure designed to attenuate spurious correlations due to volume conduction.

A complete list of the 23 biomarkers across the 3 families is provided in Supplementary Table S2.

#### 9.6.2 Spatial and temporal aggregation

For each biomarker *B*, values were first computed at the level of individual channels and epochs. To obtain center- agnostic representations, two complementary reductions were applied:

**Figure S6:**
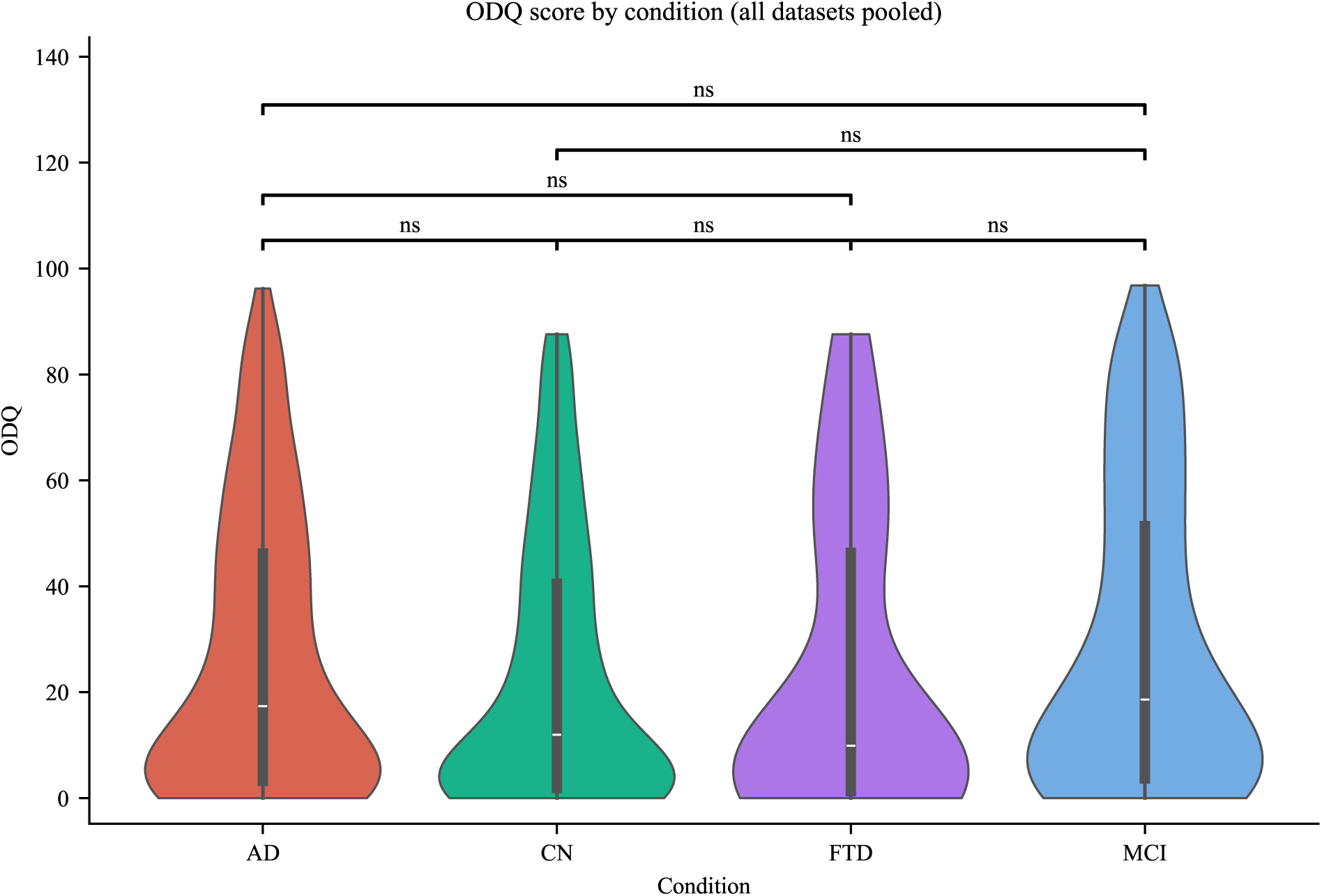
Distribution of the Overall Data Quality (ODQ) score across diagnostic conditions across all centers: Each violin shows the full distribution of ODQ scores; Pairwise significance labels reflect Mann-Whitney, “ns” denotes no significant difference. No condition pair showed a statistically significant difference in ODQ.

- Spatial summaries across channels
- **–** Channel mean: reflects overall magnitude of the marker on the scalp.
- **–** Channel standard deviation: reflects **global field power**, i.e., the spatial heterogeneity of the marker across electrodes.
- Temporal summaries across epochs
- **–** 80% trimmed mean: captures typical activity while reducing sensitivity to residual artifacts.
- **–** Standard deviation: quantifies **temporal variability** of the marker within the recording.

Combining these dimensions yielded four descriptors for each biomarker:

- *µ/µ*: mean across channels and trimmed mean across epochs (average activity),
- *ó/µ*: standard deviation across channels and trimmed mean across epochs (global field power),
- *µ/ó*: mean across channels and standard deviation across epochs (temporal variance),
- *ó/σ*: standard deviation across channels and epochs (joint spatiotemporal variability).

This dual aggregation produced **92 feature subtypes** (23 biomarkers × 4 descriptors) used as input to the classification models.

#### 9.6.3 Implementation

All computations were performed with fixed parameters across centers. Spectral, information-theoretic, and connectivity features were extracted from identical epochs defined in the preprocessing stage. No feature selection or parameter tuning was performed prior to model training to preserve independence between feature extraction and classification.

**Figure S7:**
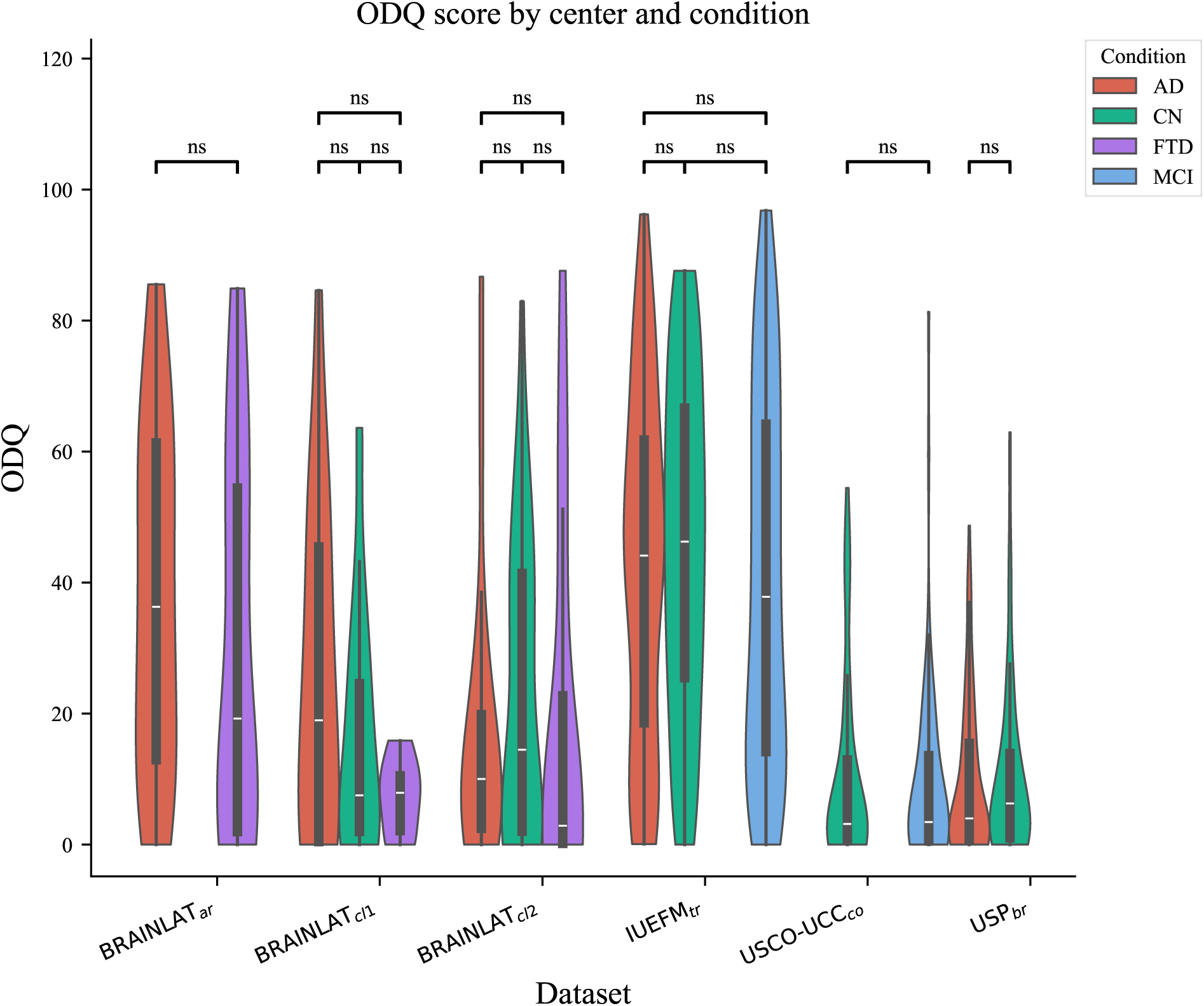
Distribution of the Overall Data Quality (ODQ) score stratified by center and diagnostic condition: Each x position corresponds to a specific center, and violins are colored by diagnostic conditions inside each center. Pairwise significance brackets reflect Mann-Whitney U tests between condition within each center; “ns” denotes no significant difference. No center exhibited a significant ODQ difference across diagnostic groups.

### 9.7 Extended evaluation, calibration, and brain–behavior association analyses

Model performance across all validation folds was evaluated using the area under the receiver operating characteristic curve (ROC-AUC), a threshold-independent metric quantifying the discriminative ability between diagnostic categories.

To assess the reliability of the predicted probabilities, we compared the uncalibrated and calibrated model outputs (Figure 1 Calibration) to determine i) whether the model was intrinsically well-calibrated (*Cal_err_*) and ii) whether calibration procedures improved the alignment between predicted and true outcome frequencies (*Cal*_Δ_). Model calibration was performed using CalibratedClassifierCV Python class provided by the scikit-learn library with Platt scaling (sigmoid calibration) and five-fold cross-validation. This procedure fits a sigmoid function to the model’s decision scores, transforming them into probability estimates that more closely reflect empirical likelihoods. For instance, in a well-calibrated model, among all cases predicted with a 60% probability of being MCI, approximately 60% should actually belong to that category.

Calibration, however, cannot compensate for a poorly discriminative classifier: if the base model fails to separate diagnostic groups, no post-hoc transformation can yield meaningful probabilities. Furthermore, even for accurate models, calibration may remain imperfect when the decision function is highly non-monotonic or when score distributions are discrete, as often occurs in tree-based ensembles such as Random Forests.

To assess whether the observed classification performance exceeded chance level, we implemented a two-stage statistical validation framework (Figure 1 Permutation Test).

Stage 1 — Repeated cross-validation. To enhance robustness, we conducted 500 repetitions of k-fold cross-validation. At each repetition, the dataset was randomly partitioned into new folds, and model predictions were obtained exclusively from out-of-fold test samples, ensuring independence between training and testing data. This procedure yielded a distribution of 500 AUC values for each validation strategy, classification task and center, representing the variability and stability of model performance across split iterations.

Stage 2 — Permutation testing. To evaluate statistical significance, we performed 500 label permutations per task using the same cross-validation structure. In each permutation, diagnostic labels were randomly shuffled while preserving data structure and class proportions, thereby generating a null distribution of performance metrics expected under the assumption of no true association between EEG features and diagnostic categories.

Statistical inference (Figure 1 Performance Distributions). Model performance was then assessed using the difference of means between the true labels AUCs and the permuted labels AUCs. We computed the significance level using the proportion of permuted AUCs greater than the observed median of real AUC as the empirical *p_val_*, thus estimating the probability that a model trained on data with no true association between markers and diagnostic categories could achieve comparable performance. Finally, we corrected the *p_val_* wit FDR using the Benjamini–Hochberg procedure across all obtained *p_val_*.

To quantify the consistency of individual-level model predictions across validation strategies, we analyzed correlations between predicted probabilities obtained under within-center, multi-center, and cross-center validation. For each diagnostic contrast, predictions were extracted exclusively from out-of-fold test samples to avoid information leakage. Subject-level predicted probabilities were averaged across repetitions prior to correlation analysis.

Spearman rank correlations were computed between each pair of validation strategies (within vs multi, within vs cross, and multi vs cross) to assess the stability of subject ranking across training conditions. Correlations were evaluated separately for each diagnostic contrast and, where applicable, within individual centers. Statistical significance was assessed using the *p_val_* obtained from the Spearman rank statistic, and FDR correction using the Benjamini–Hochberg procedure was applied across all correlations to account for multiple comparisons.

Associations between model predictions and cognitive measures were computed using residualized Spearman correlations. For each subject, predicted probabilities were extracted from out-of-fold test predictions under within-center, multi-center, or cross-center validation and averaged across repeated cross-validation iterations when applicable. cogni- tive measures included measures of global cognition, memory, language, attention, and executive function, depending on availability at each center.

To control for demographic confounds, both predicted probabilities and cognitive scores were independently residualized with respect to age, sex, and years of education using linear regression and Spearman rank correlations were computed between residuals using the python package *Pingouin* (Vallat, 2018), which comes with a build-in function to do this (partial_corr). Analyses were conducted separately for each diagnostic contrast and validation strategy. Statistical significance was evaluated with FDR correction using the Benjamini–Hochberg procedure per validation strategy, cognitive measures and contrasts. In the correlation heatmaps, all coefficients are displayed, with color indicating correlations that survived FDR correction.

Finally, to identify the EEG features most discriminative across diagnostic groups and to provide interpretable insights into model decisions, we applied SHAP (SHapley Additive exPlanations) analysis (S. Lundberg & Lee, 2017; S. M. Lundberg et al., 2020) (Figure 1 SHAP Values Feature Importance). SHAP values quantify the magnitude and direction of each feature’s contribution to individual predictions, thereby enabling both global feature importance ranking and local interpretability. This approach enhances transparency by revealing which EEG characteristics most strongly influence diagnostic classification.

To visualize the effect of individual markers, we examined the ten features with the highest absolute mean SHAP values for each classification task, analyzing how their magnitude related to SHAP contribution—that is, whether higher or lower feature values increased the likelihood of predicting one class over another. Subsequently, the five most influential features per task where all pooled together in a radar plot to facilitate cross-task comparison.

**Table S2:**
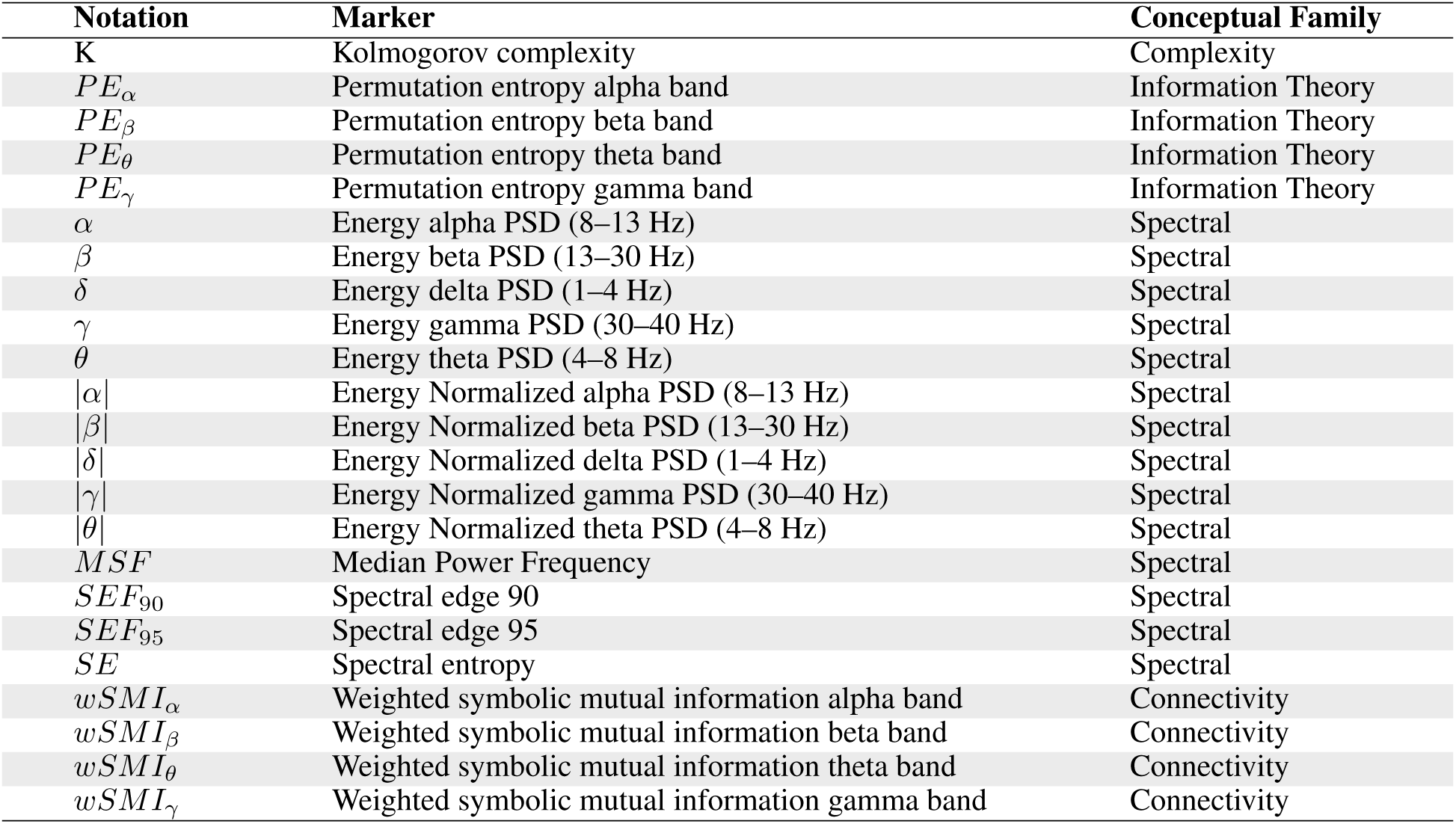
Summary of the EEG markers considered in the study taken from (Sitt et al., 2014; Engemann et al., 2018). The notation column shows the abbreviated symbol used in analyses, the marker column provides the full description (including frequency ranges for spectral measures), and the conceptual family indicates the theoretical category of each marker.

**Table S3:** Subject level predicted probabilities correlations between validation strategies by diagnostic contrast and Center.

| Contrast | Center | Correlation | r | p-value |
| --- | --- | --- | --- | --- |
| CN vs AD | $BRAINLAT_{cl1}$ | Multi vs Cross | 0.94 | 0.00 |
|  |  | Within vs Cross | 0.68 | 0.00 |
|  |  | Within vs Multi | 0.83 | 0.00 |
| | $IUEFM_{tr}$ | Multi vs Cross | 0.89 | 0.00 |
|  |  | Within vs Cross | 0.76 | 0.00 |
|  |  | Within vs Multi | 0.95 | 0.00 |
| | $BRAINLAT_{cl2}$ | Multi vs Cross | 0.96 | 0.00 |
|  |  | Within vs Cross | 0.80 | 0.00 |
|  |  | Within vs Multi | 0.88 | 0.00 |
| | $USP_{br}$ | Multi vs Cross | 0.93 | 0.00 |
|  |  | Within vs Cross | 0.83 | 0.00 |
|  |  | Within vs Multi | 0.95 | 0.00 |
| CN vs MCI | $IUEFM_{tr}$ | Multi vs Cross | 0.33 | 0.00 |
|  |  | Within vs Cross | 0.17 | 0.02 |
|  |  | Within vs Multi | 0.97 | 0.00 |
| | $USCO - UCC_{co}$ | Multi vs Cross | 0.44 | 0.00 |
|  |  | Within vs Cross | -0.01 | 0.89 |
|  |  | Within vs Multi | 0.76 | 0.00 |
| FTD vs AD | $BRAINLAT_{ar}$ | Multi vs Cross | 0.69 | 0.00 |
|  |  | Within vs Cross | -0.05 | 0.78 |
|  |  | Within vs Multi | 0.62 | 0.00 |
| | $BRAINLAT_{cl2}$ | Multi vs Cross | 0.13 | 0.38 |
|  |  | Within vs Cross | -0.24 | 0.09 |
|  |  | Within vs Multi | 0.88 | 0.00 |

